# Exercise Training Outcomes in Patients with Chronic Heart Failure with Reduced Ejection Fraction Depend on Medications and Disease Conditions: Proposal of SEEM Score to Judge Exercise Suitability

**DOI:** 10.1101/2022.02.26.22271542

**Authors:** Yukako Soejima, Hideki Yoshioka, Sayuri Guro, Hiromi Sato, Hiroto Hatakeyama, Yasunori Sato, Yoshihide Fujimoto, Naohiko Anzai, Akihiro Hisaka

## Abstract

**Background:** Exercise training is an accepted evidence-based adjunct treatment modality for patients with chronic heart failure. However, the influence of medications or conditions on exercise has not been fully explored.

**Methods:** The patient records of the Heart Failure: A Controlled Trial Investigating Outcomes of Exercise Training (HF-ACTION) study were obtained from the National Heart, Lung, and Blood Institute, and analyzed by medications used at baseline (renin-angiotensin system inhibitors [RASIs], beta-blockers [BBs], and combination of both) with multivariable Cox regression models focusing on the interactions with exercise effects, and a score to indicate exercise training suitability was proposed accordingly.

**Results:** Medication type strongly influenced the exercise effect on all-cause death (AD) (P = 0.007) even though medication itself did not change prognosis significantly in HF-ACTION trial. In patients taking both BBs and RASIs at baseline, exercise reduced the AD risk (hazard ratio [HR], 0.86; 95% confidence interval [CI], 0.75–0.98), whereas in patients not taking BBs, exercise increased the risk (HR, 4.58; 95% CI, 2.90–6.86). The exercise on AD was also influenced by pulse pressure, hemoglobin level, electrocardiography conditions, body mass index, and history of stroke. Accordingly, we constructed the Score for Eligibility of Exercise on Mortality (SEEM). When exercise training was chosen based on SEEM score, both AD and AD and hospitalization (ADH) risks were expected to reduce significantly (HR, 0.54; 95% CI, 0.44–0.68; HR, 0.83; 95% CI, 0.75–0.93, respectively).

**Conclusions:** Exercise training in patients with heart failure should be recommended carefully with consideration of patient background.

**Clinical Trial Registration:** Clinicaltrials.gov identifier: NCT00047437. Trial registration date: 4 October 2002.

## BACKGROUND

Exercise therapy improves mortality, health-related quality of life, and exercise capacity in patients with chronic heart failure^**1–6**^ and is widely recommended as an effective treatment that can be combined with pharmacotherapy such as renin-angiotensin system inhibitors (RASIs, i.e., angiotensin-converting enzyme inhibitor and/or angiotensin II receptor blocker) and beta-blockers (BBs).^**7,8**^ The Heart Failure: A Controlled Trial Investigating Outcomes of Exercise Training (HF-ACTION) study conducted in 2003–2008 is the only large-scale randomized controlled trial that examined the efficacy of exercise therapy in medically stable patients with chronic heart failure.^**9**^ The trial demonstrated that a prescribed exercise training program was associated with a reduction in all-cause death and hospitalization (ADH) rate. This improvement was statistically significant after the adjustment for prognostic factors, and a non-significant reduction in all-cause death (AD), one of the secondary endpoints, was noted regardless of factor adjustment.

In chronic heart failure, reducing the cardiac load is essential for successful treatment.^**10–12**^ We previously performed a model-based meta-analysis of 61 studies of patients with chronic heart failure and reported that estimated myocardial oxygen consumption, a cardiac load index, correlated excellently with the reduction in mortality after various pharmacotherapies.^**13**^ In particular, RASIs and BBs reduced the estimated myocardial oxygen consumption more efficiently than other drugs, including calcium channel blockers and direct renin inhibitors, which supported the superior prognostic improvement ability of these drugs. In contrast, exercise activates the sympathetic nervous system^**14**^ and increases cardiac load, which contradicts the expected effect of drug therapy.^**15–17**^ Therefore, the use of RASIs and BBs may affect exercise training efficacy; however, no studies have examined the interaction between medication and exercise. Although the subgroup analysis of the HF-ACTION trial showed that baseline RASI and BB use did not affect exercise effects, there was no quantitative analysis of the extent to which exercise effects differed by particular medication combination group. Additionally, the benefits of exercise training are potentially influenced by the patient’s cardiopulmonary capacity, medical condition, and medical history.^**18–20**^

This study aimed to quantitatively reveal the impact of baseline medication use/various patient conditions on exercise training effects and provide evidence for identifying patients who would benefit from or be harmed by exercise training. This was a post hoc analysis of individual patient data from the HF-ACTION trial. Interactions with exercise that were statistically significant for the outcomes compared with the overall average were systematically explored using Cox proportional hazards models. Accordingly, the Score for Eligibility for Exercise on Mortality (SEEM) score was constructed to determine exercise training suitability.

## METHODS

Here we analyzed anonymized individual patient-level data from the HF-ACTION trial obtained from the National Heart, Lung, and Blood Institute, USA. The study protocol and design were reviewed and approved by the ethical review board of the Graduate School of Pharmaceutical Sciences at Chiba University.

### Study Population

The HF-ACTION study was a randomized multicenter trial that evaluated the effectiveness of exercise training versus usual care in patients with chronic heart failure. The study included patients with stable heart failure with a left ventricular ejection fraction (LVEF) of less than 35% and a New York Heart Association (NYHA) class of II– IV despite optimal heart failure treatment for at least 6 weeks. The details are described in the original report for HF-ACTION study.^**9**^

### Outcomes

The primary endpoints in this analysis were (1) AD; and (2) ADH during the entire follow-up period (up to 4 years; median, 30 months). We adopted an intention-to-treat population analysis because the study purpose was to explore factors to discriminate the benefit of exercise training, including patient exercise tolerance.

### Statistical Analysis

#### Stratification by Medication at Baseline

Patients were categorized into four groups according to the use of BB and RASIs at baseline: both BB and RASI, RASI only, BB only, and neither BB nor RASI. In this study, angiotensin-converting enzyme inhibitors (ACEIs) and angiotensin II receptor blockers (ARBs) were combined as RASIs owing to their similar mechanisms of action. Heterogeneity in patient characteristics between these groups was tested using ANOVA or the Kruskal-Wallis test for continuous variables and the Chi-square test for categorical variables. The comparison was not performed for the subgroup of patients who were not treated with RASI or BB at baseline because the sample size was too small (n = 10; 0.5%). For comparative purpose with the Cox proportional hazard analysis, the cumulative rates of events in each group were analyzed using the Kaplan-Meier method and the log-rank test.

#### Cox Proportional Hazards Models

All available baseline characteristics were extracted from the data as candidates (**Supplementary Table S1**). For continuous variables, missing variables with a rate of < 30% were complemented by a multiple imputation method and then included. Spearman’s rank correlation coefficient of greater than 0.8 for continuous variables and the chi-square value of greater than 300 for categorical variables were excluded. All continuous variables were transformed into binary variables nearly at the 25th, 50th, and 75th percentiles. For each AD and ADH, using Cox proportional hazards models, significant prognostic factors and their interactions with exercise were selected from 73 candidates (**Supplementary Table S1**) with the stepwise methods and confirmed cautiously by the bootstrap method. In these processes, the use or not of RASIs and/or BBs at baseline was always included in the model. Finally, the association between the baseline medication and clinical outcomes was evaluated from the model. Since the number of patients who were not taking both RASI and BB at baseline was extremely small (n = 10 [0.5%]), this group was not analyzed for the exercise effect. The details are provided in the **Supplementary Statistical Methods**.

#### Scores Predicting Benefit or Harm from Exercise Training

Based on the results of the Cox proportional hazards model, the SEEM score was developed incorporating significant interaction terms identified in the final model. For ease of use, all predictors comprising the score were assigned integer points by scaling of the β-coefficients of the final Cox model. The score was designed such that the higher the total score, the more beneficial exercise training would be. That is, given a cutoff value, if a patient’s score was equal to or greater than it, exercise training would be recommended.

#### Software

The PHREG procedure implemented in SAS® 9.4 was used to conduct the Cox proportional hazards model analysis. Python 3.8 (Python Software Foundation) was used for the data preprocessing and illustrations. IterativeImputer, implemented in scikit-learn 0.24.2, was used for multiple imputations of missing data.

## RESULTS

### Patient Characteristics

All eligible patient records (2130 of 2331 patients consented to share for non-commercial uses and allowed for any research purpose) were included in the analysis. Clinical outcomes and representative patient characteristics according to baseline medication status are summarized in **Table 1**. A complete table is available in the supplemental materials (**Supplementary Table S2**). Although some differences were observed in background characteristics between the medication groups, significant difference was not detected in clinical outcomes.

**Table 1.**
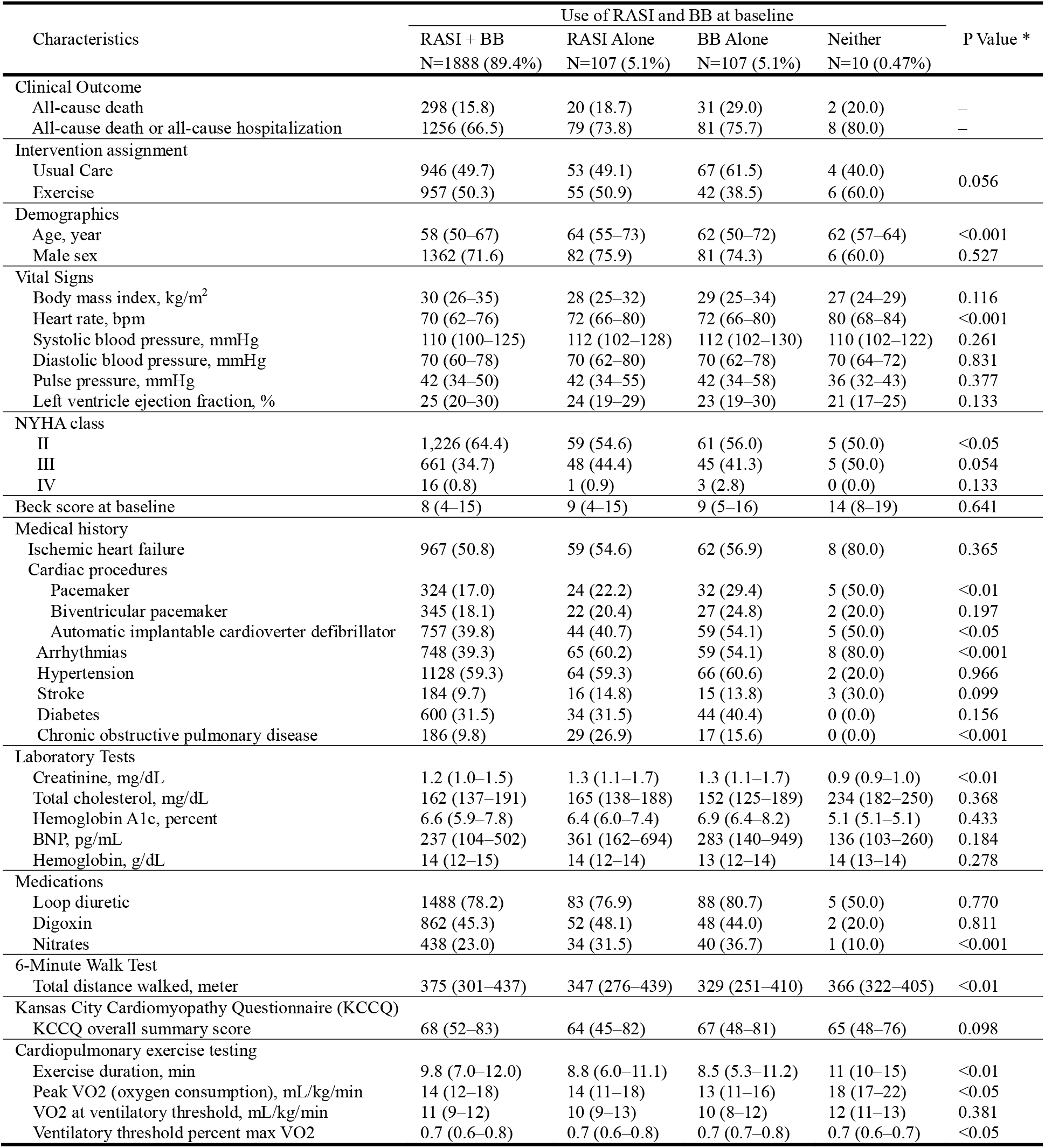
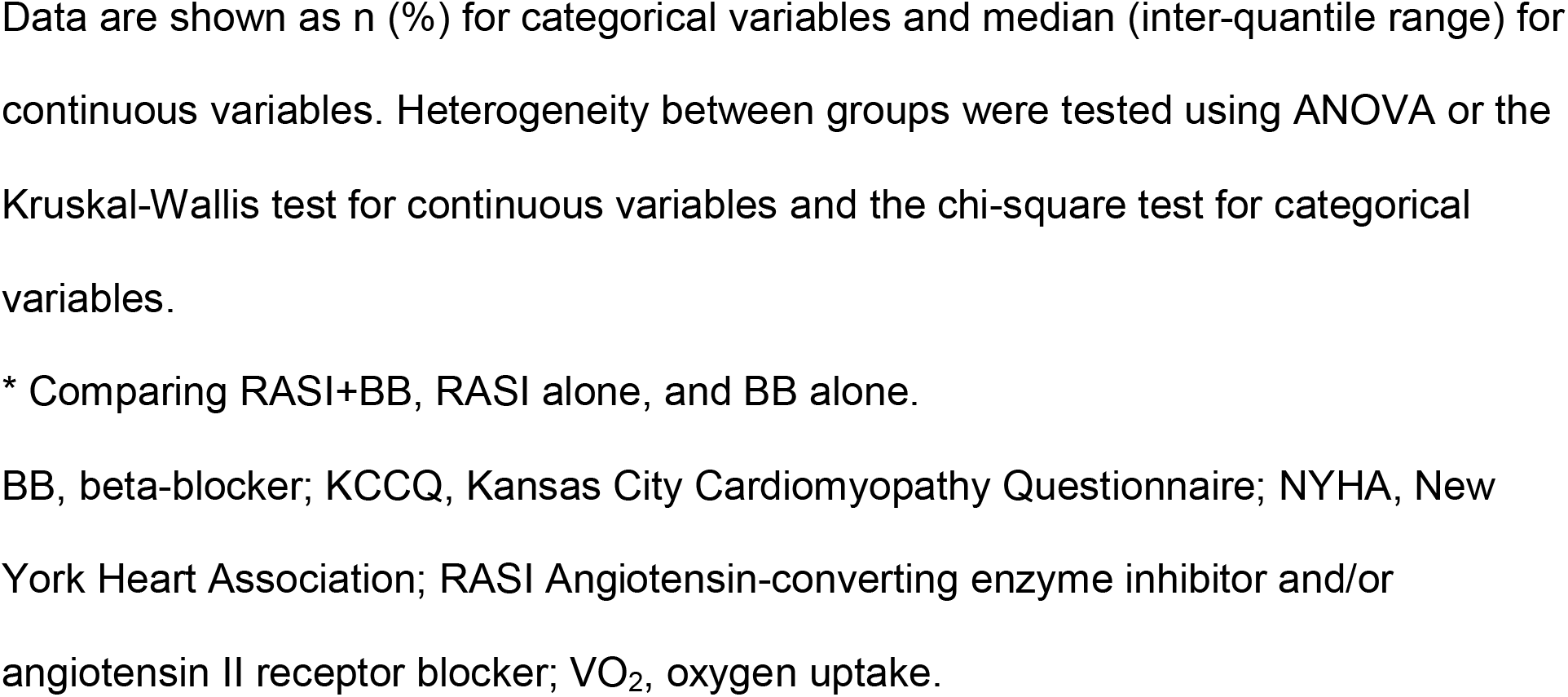
Clinical Outcomes and Patient Characteristics by Baseline RASI and BB Medication Status.

### Multivariate Cox Proportional Hazards Model Analysis

**Figures 1 and 2** summarize the influence of RASI and/or BB use at baseline and other patient characteristics on the effectiveness of exercise training using forest plots for AD and ADH, respectively. Other exercise-independent effects and parameter estimates are shown in **Supplementary Figures S1 and S2 and Supplementary Tables S3 and S4**. The hazard ratios (HRs) of the exercise effect in all patients were 0.92 (95% CI, 0.81–1.04) for AD and 0.88 (95% CI, 0.82–0.94) for ADH (**Figures 1A and 2A**), which was consistent with the original analysis of the HF-ACTION trial reporting that exercise training significantly reduced the risk of ADH with adjustment but not that of AD. The present analysis revealed that the effects of exercise on AD differed significantly depending on RASI and/or BB medication status at baseline (interaction P = 0.007). A tendency toward beneficial exercise effects was estimated in the subgroups taking both RASIs and BBs at baseline (HR, 0.86; 95% CI, 0.75–0.98) or BBs alone (HR, 0.63; 95% CI, 0.38–1.00), whereas exercise training was associated with a notably increased risk of AD in patients treated with RASIs alone at baseline (i.e. not treated with BBs, HR, 4.58; 95% CI, 2.90–6.86). In contrast, for ADH, the overall interaction between RASIs and/or BBs at baseline and the exercise effect did not reach statistical significance (interaction P = 0.119). However, exercise tended to be effective for ADH in patients treated with both RASIs and BBs (HR, 0.88; 95% CI, 0.82–0.94) and BBs alone (HR, 0.55; 95% CI, 0.39–0.78), while in patients treated with RASIs alone, the effect of exercise training was insignificant (HR, 0.97; 95% CI, 0.70-1.37).

**Figure 1.**
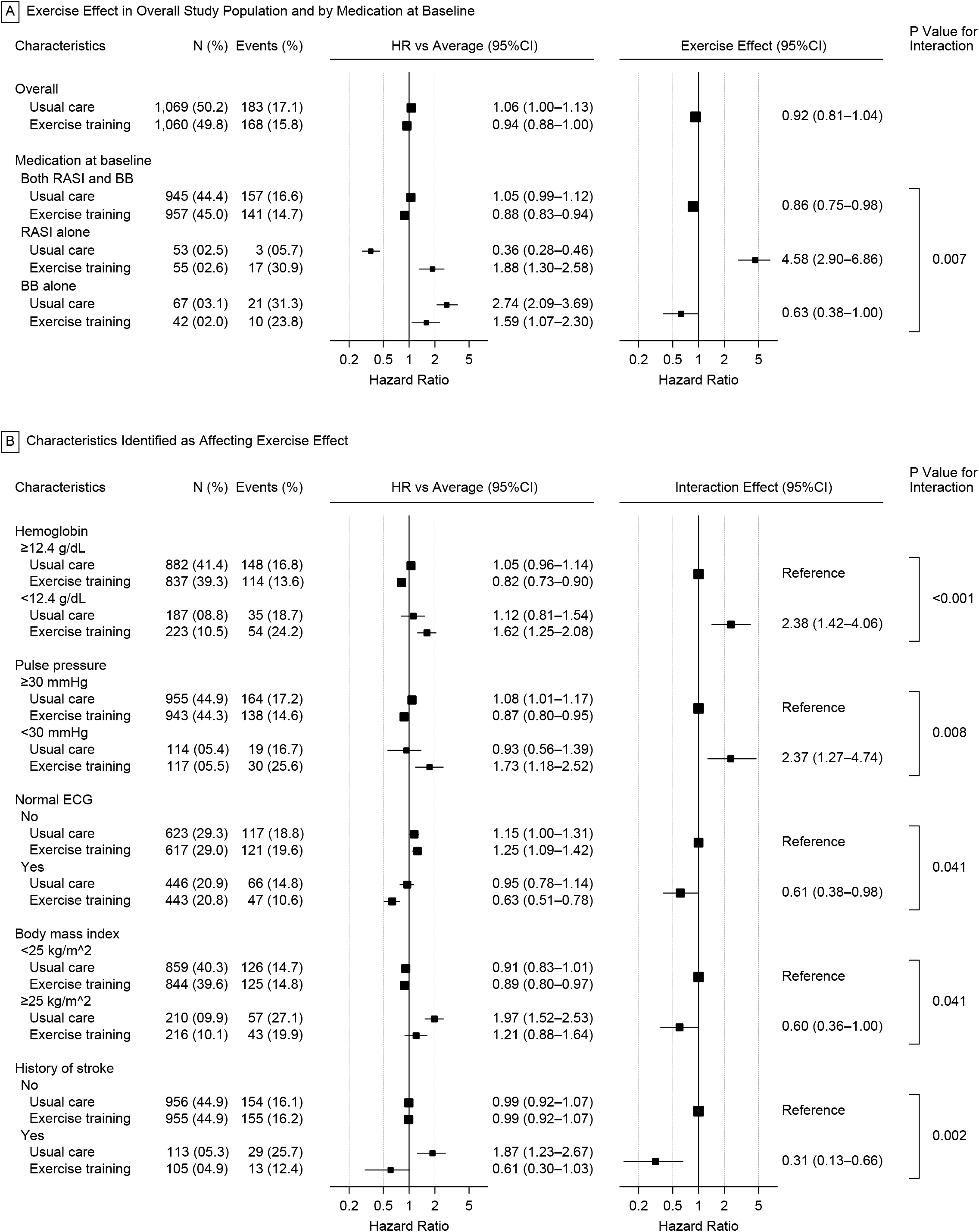
Influence of Medication and Other Characteristics on Exercise Effect for All-Cause Death All the HRs, effects and their CIs were calculated from the analysis of 1,000 bootstrap datasets based on the multivariate Cox proportional hazards model analysis. Interaction effects represent the relative effect of exercise training when the exercise effect in the reference group is assumed to be 1. The hazard ratio for a given subgroup vs the population mean was calculated by subtracting the mean of the linear predictors for all patients from the mean of the linear predictors for patients belonging to that subgroup. *BB* beta-blocker, *CI* confidence interval, *ECG* electrocardiogram, *HR* hazard ratio, *RASI* angiotensin-converting enzyme inhibitor and/or angiotensin II receptor blocker.

**Figure 2.**
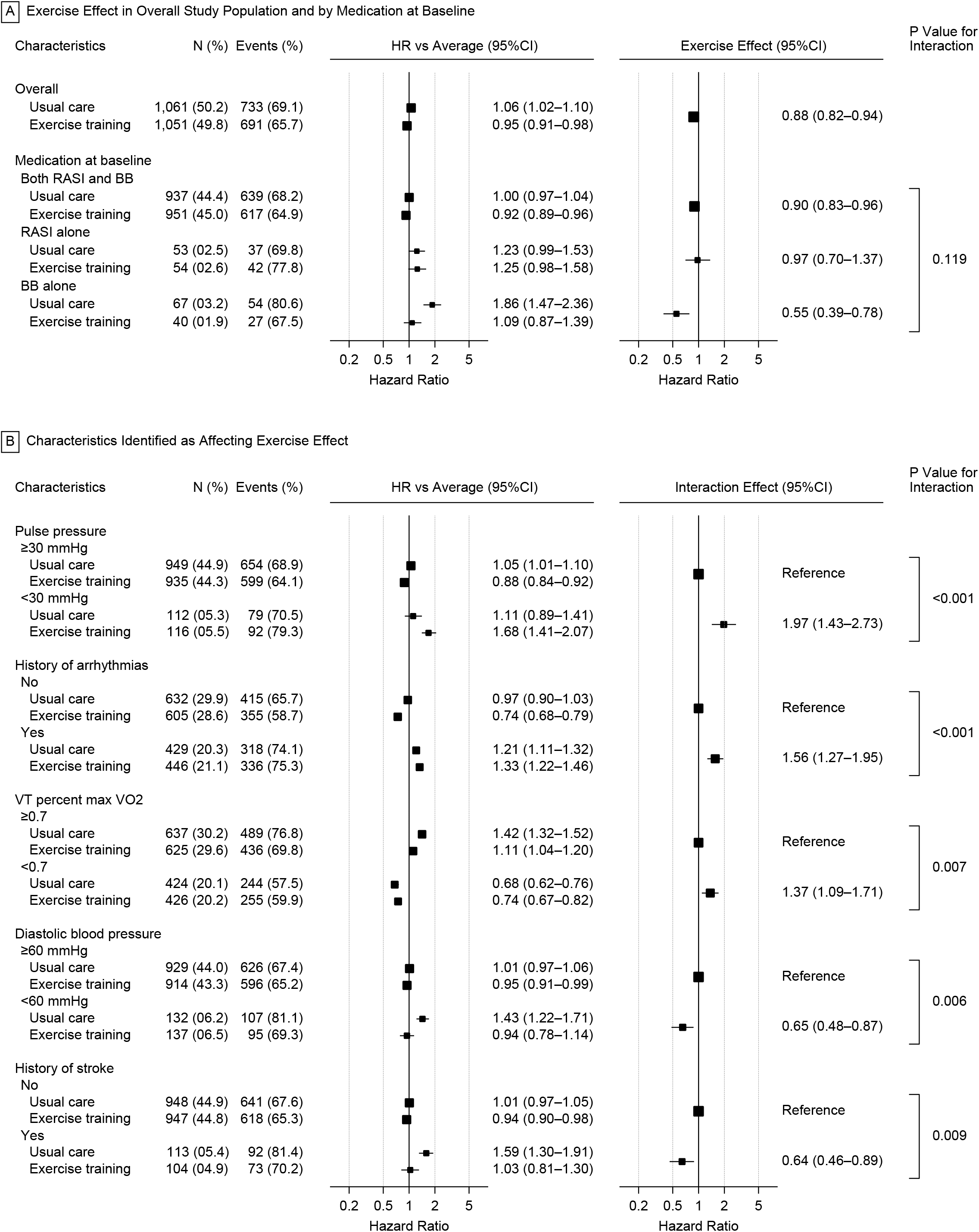
Influence of Medication and Other Characteristics on Exercise Effect for All-Cause Death or Hospitalization All the HRs, effects and their CIs were calculated from the analysis of 1,000 bootstrap datasets based on the multivariate Cox proportional hazards model analysis. The figure was prepared in the same format as Figure 1. *BB* beta-blocker, *CI* confidence interval, *ECG* electrocardiogram, *HR* hazard ratio, *RASI* angiotensin-converting enzyme inhibitor and/or angiotensin II receptor blocker, *VO*_*2*_ oxygen uptake.

For both endpoints, five factors were identified as affecting the exercise effect (**Figures 1B and 2B**). In particular, a pulse pressure ≥ 30 mmHg and a history of stroke were commonly identified as factors that enhanced the effectiveness of exercise training. Other factors identified as reducing exercise effectiveness included a hemoglobin of < 12.4 g/dL for AD versus a VT percent max VO_2_ of < 0.7 and a history of arrhythmia for ADH, while those identified as enhancing exercise effectiveness included a normal electrocardiography (ECG) findings and a body mass index (BMI) of < 25 kg/m^2^ for AD versus a diastolic blood pressure of < 60 mmHg for ADH. The baseline medication status was not a significant prognostic factor for either AD or ADH, and the linear predictors calculated from the main effects other than the medication were almost the same (**Supplementary Figure S3**).

To confirm the consistency between this study and previous analyses of the HF-ACTION study, we also performed Cox proportional hazards model analyses in which the interactions of medication groups or all interactions were excluded from the final model (**Supplementary Tables S5 and S6**). The estimated coefficients and significance of the main effects did not change with the absence or presence of interactions with exercise.

The cumulative incidence curves for AD and ADH according to baseline RASI and/or BB medication status are shown in **Figures 3 and 4**, respectively. The results showed a similar trend to the results of the Cox regression analysis in all medication groups. In patient groups taking RASIs or BBs alone, the HRs of the exercise group for AD were increased more than 1 year later after the exercise therapy began. This may have been possibly caused by a change in medication during the trial. Other possibilities include considerably delayed appearance of harmful effects by exercise. A subgroup analysis was performed stratifying by the presence of a medication change from baseline but failed to explain the observed HR changes (**Supplementary Figure S4 and S5**).

**Figure 3.**
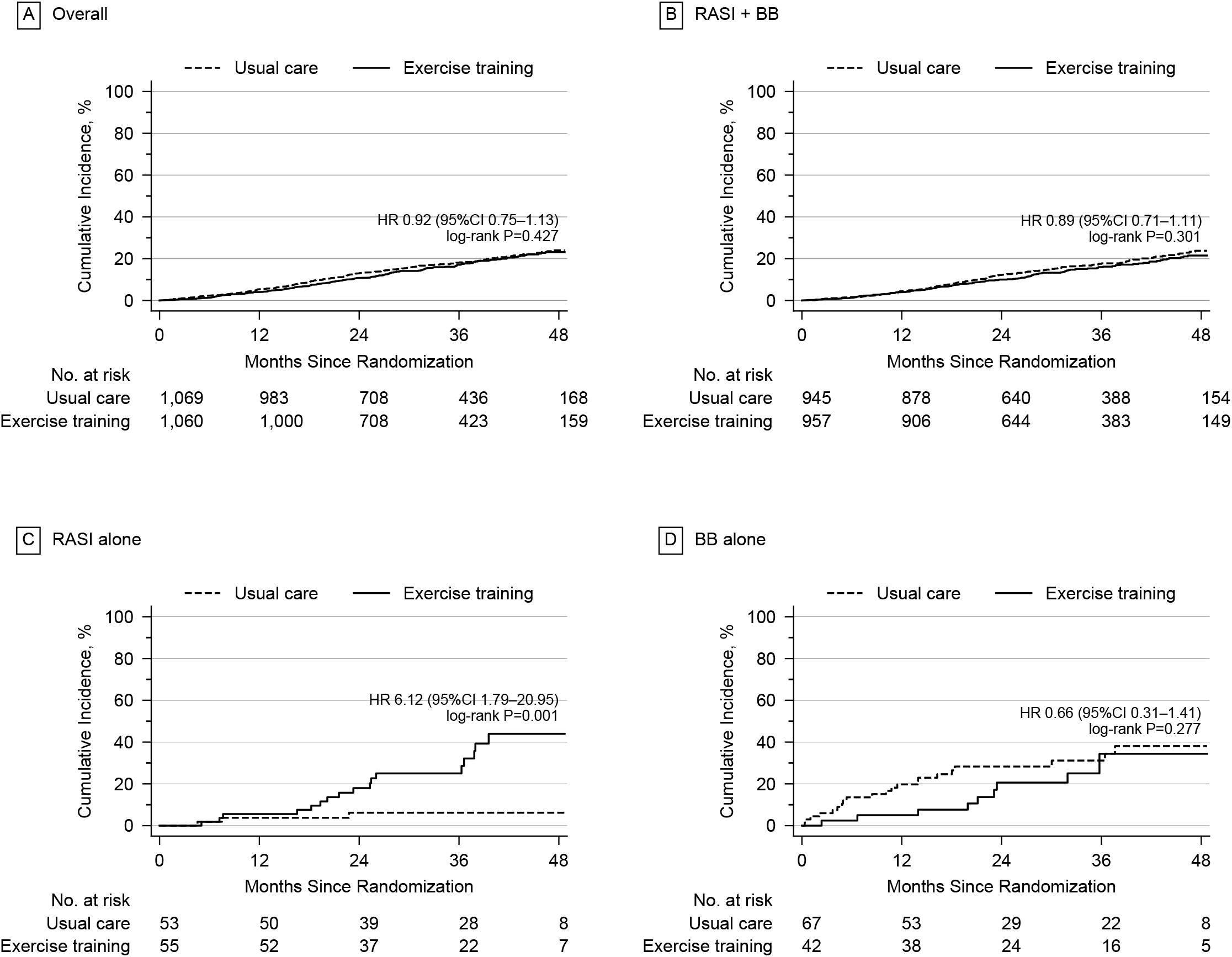
Cumulative Incidence of All-Cause Death by Medication at Baseline The cumulative incidence of all-cause death in each medication group was directly compared. **A** Overall study population. **B** Patients taking both RASIs and BBs at baseline. **C** Patients taking RASIs alone at baseline. **D** Patients taking BBs alone at baseline. *BBs* beta-blockers, *CI* confidence interval, *HR* hazard ratio, *RASIs* angiotensin-converting enzyme inhibitors and/or angiotensin II receptor blockers.

**Figure 4.**
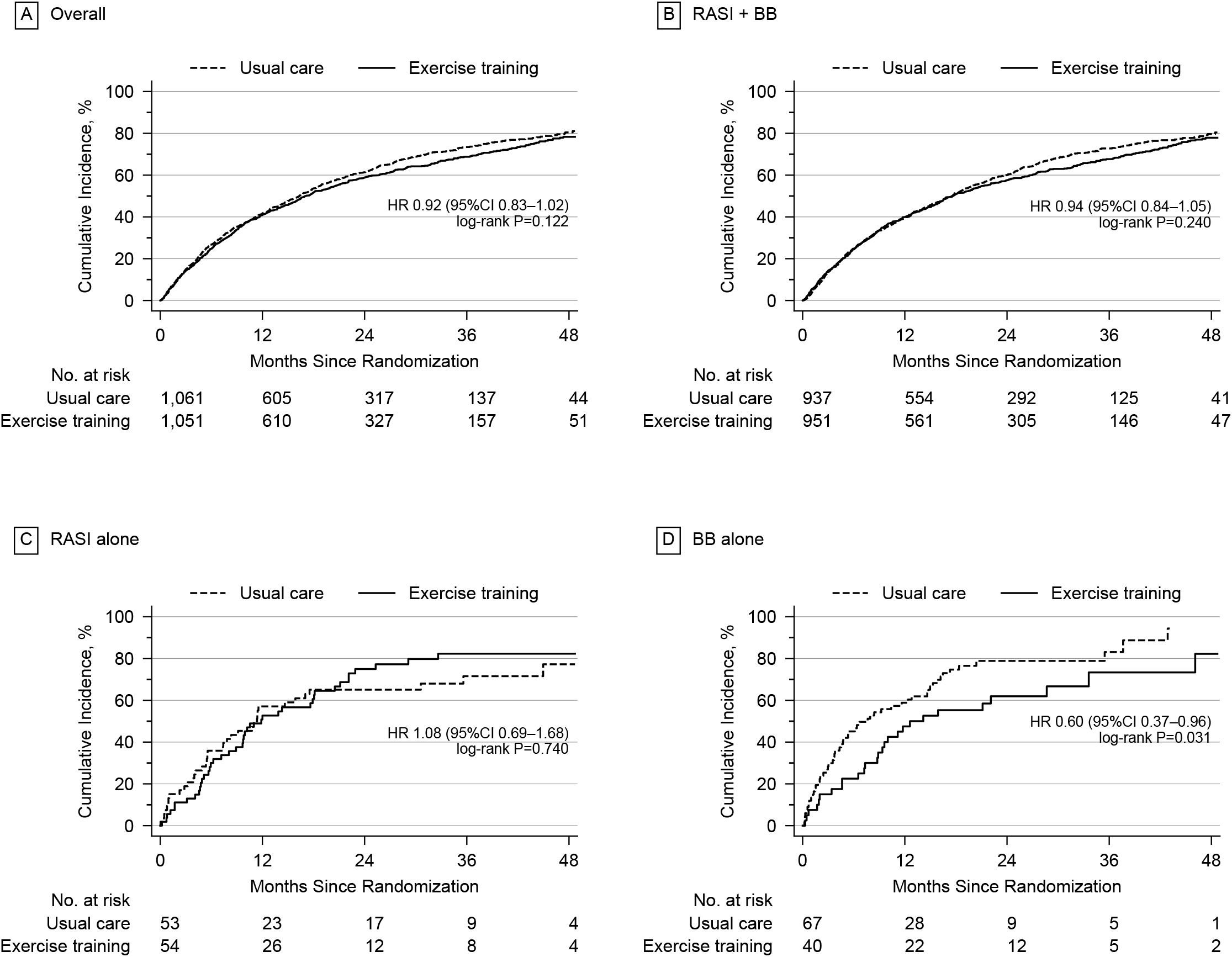
Cumulative Incidence of All-Cause Death or Hospitalization by Medication at Baseline The cumulative incidence of all-cause death or hospitalization for each medication group was directly compared. **A** Overall study population. **B** Patients taking both RASIs and BBs at baseline. **C** Patients taking RASIs alone at baseline. **D** Patients taking BBs alone at baseline. *BBs* beta-blockers, *CI* confidence interval, *HR* hazard ratio, *RASIs* angiotensin-converting enzyme inhibitors and/or angiotensin II receptor blockers.

### Score for Predicting Exercise Eligibility

To make the obtained models usable in clinical settings, we developed the SEEM clinical score, which predicts a patient’s eligibility for exercise training (**Figure 5**). The SEEM comprises a total of six predictors: pulse pressure ≥ 30 mmHg (+2), history of stroke (+2), hemoglobin ≥ 12.4 g/dL (+2), BMI ≥ 25 kg/m^2^ (+1), normal ECG findings (+1), and treatment with BBs (+3). The total score ranges from 0 to 11, with an optimized cutoff value of 7 (i.e., exercise training is recommended only if a patient has a total score of 7 or higher; **Figure 6A**). Note that although we finally used the results for AD to develop the SEEM (i.e. AD model), a score developed from the results for ADH (ADH model) was also considered in a preliminary analysis (**Supplementary Figure S6**). AD model was adopted as SEEM because the overall discriminability of the score was better than that from the ADH model (**Figure 6A and 6B**).

**Figure 5.**
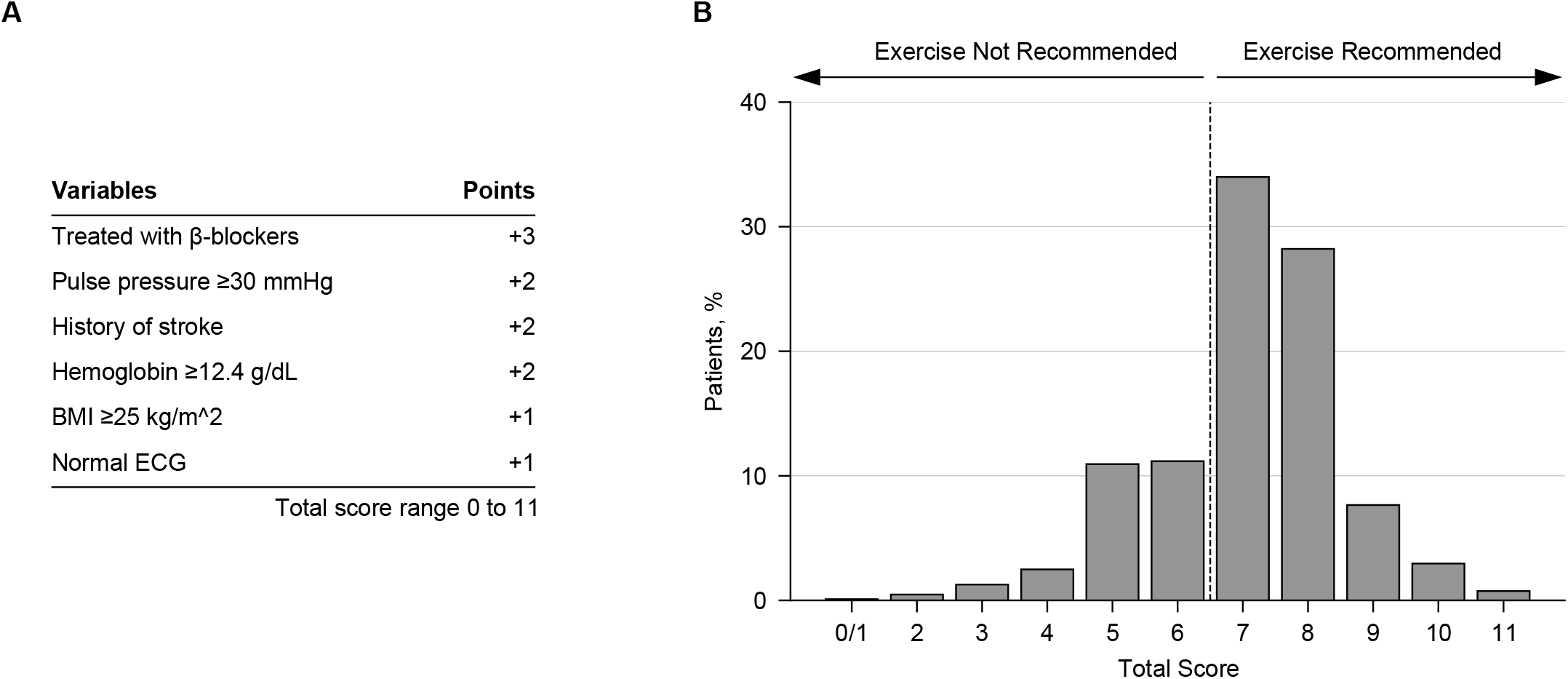
Summary of SEEM (Score for Eligibility of Exercise on Mortality) for Chronic Heart Failure Variables for points of SEEM score (**A**), and patient score distribution in HF-ACTION study (**B**). The scores incorporated all factors included in the interaction terms with exercise in the final model of the Cox proportional hazards model as well as medication groups for which the exercise effect was estimated to be significantly different from that in the overall study population. For ease of use, all variables were assigned integer points by scaling the β coefficients while maintaining the gradient of the effects. *BMI* body mass index, *ECG* electrocardiogram, *SEEM* Score for Eligibility of Exercise on Mortality.

**Figure 6.**
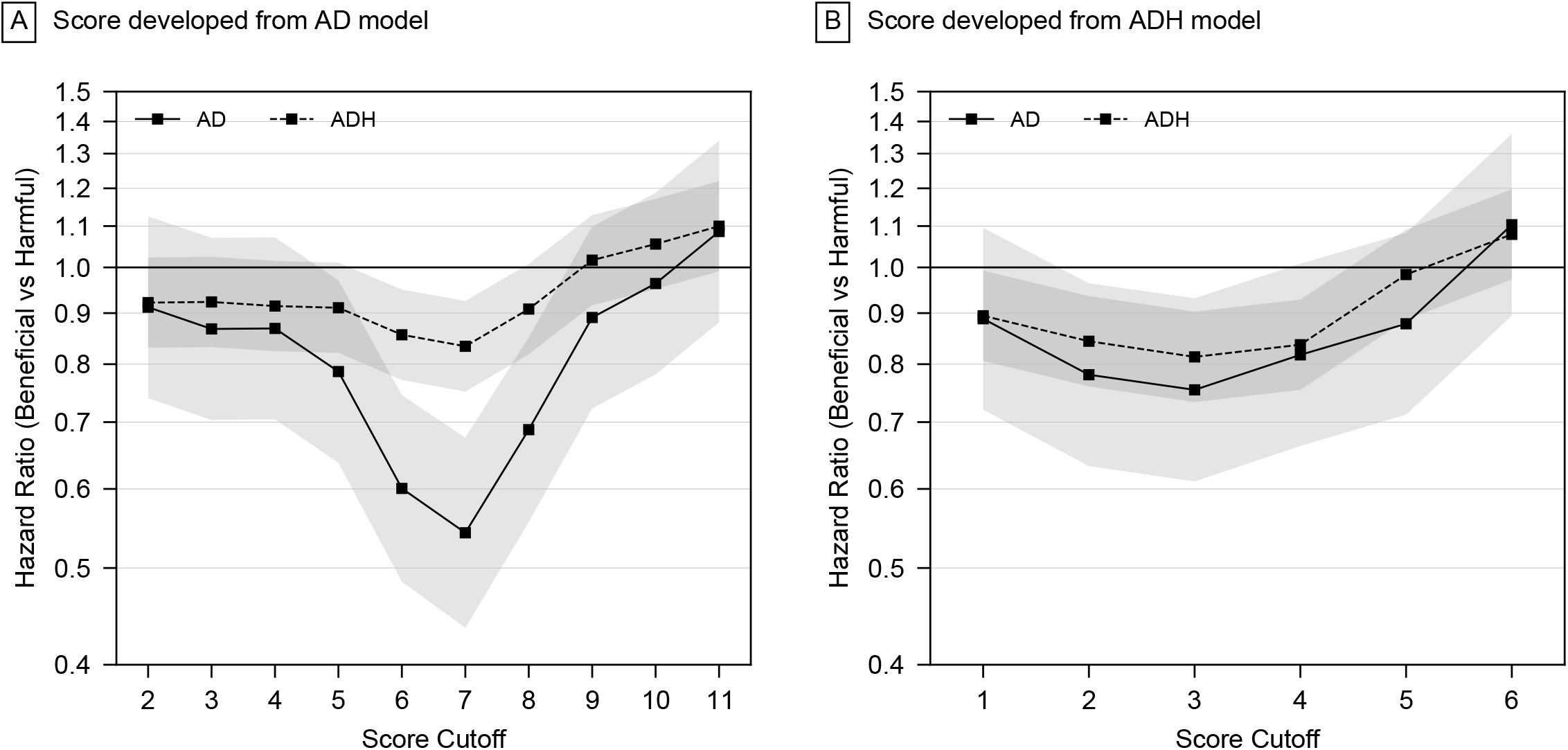
Sensitivity analysis of score cutoff for models developed by different endpoints “Beneficial” represents patients who are expected to benefit from the allocation of HF-ACTION (i.e., those in the exercise training group with a total score equal to or above the cutoff + those in the usual care group with a total score below the cutoff). “Harmful” represents patients who were predicted to have been harmed by the trial allocation (i.e., those in the usual care group with a total score equal to or above the cutoff + those in the exercise training group with a total score below the cutoff). Shaded areas represent 95% confidence intervals. *AD* all-cause death, *ADH* all-cause hospitalization.

**Figure 7** compares the cumulative incidence curves for AD and ADH between the overall study population and the subgroups for which the HF-ACTION trial allocation was expected to be beneficial (allocation beneficial) or harmful (allocation harmful) by the SEEM. Patients predicted to have benefited from the study allocation showed a markedly better prognosis for both AD and ADH, with HRs of 0.54 (95% CI, 0.44–0.68) and 0.83 (95% CI, 0.75–0.93), respectively, than those predicted to have been harmed by the study allocation. In addition, compared with the overall study population, the beneficial allocation group had a significantly lower rate of AD (HR, 0.70; 95% CI, 0.57– 0.86), indicating that, compared to a random decision, determining which patients should versus should not undergo exercise training according to SEEM score is expected to result in an additional risk reduction of 30%.

**Figure 7.**
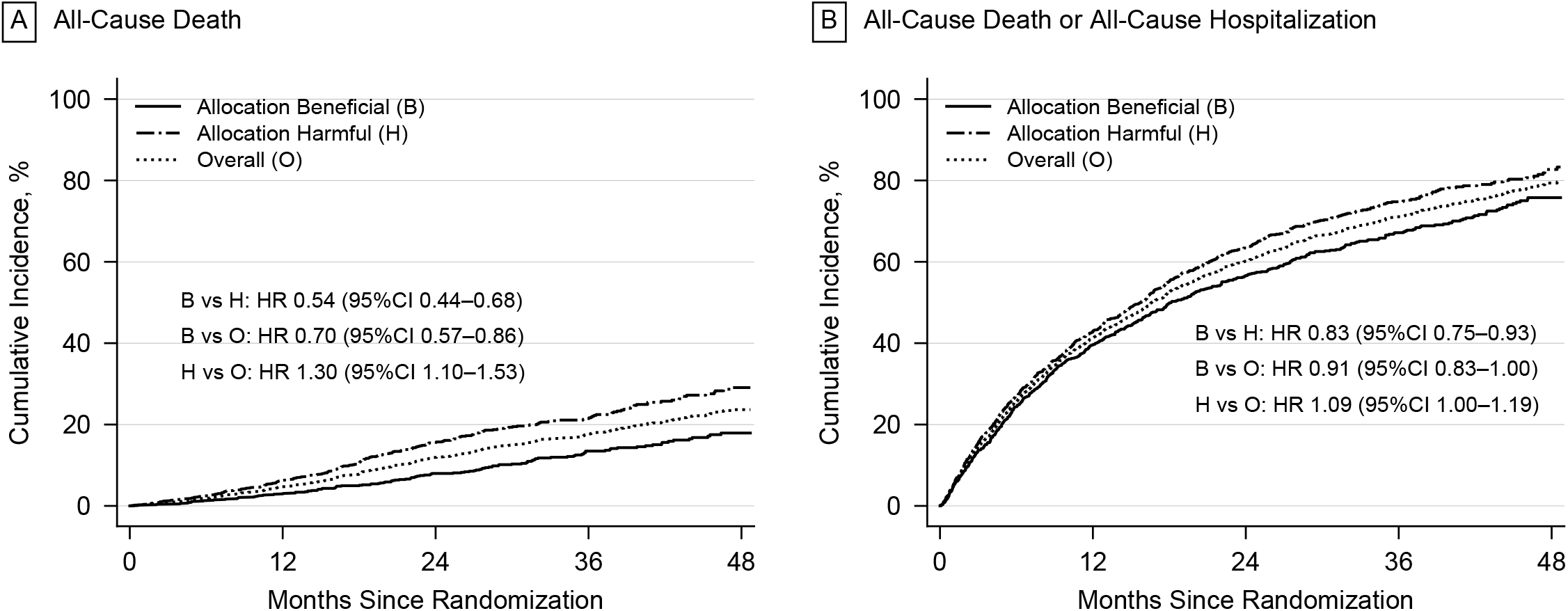
Cumulative incidence for endpoints in patients whose SEEM scores were harmful or beneficial Beneficial allocation represents patients who are expected to benefit from the allocation of HF-ACTION (i.e., those in the exercise training group with a total score equal to or above the cutoff + those in the usual care group with a total score below the cutoff). Harmful allocation represents patients who were predicted to have been harmed by the trial allocation (i.e., those in the usual care group with a total score equal to or above the cutoff + those in the exercise training group with a total score below the cutoff). The cutoff value was set to 7. *CI* confidence interval, *HR* hazard ratio.

Adherence to SEEM resulted in beneficial outcomes for both AD and ADH (**Supplementary Figure S7**) even when patients were classified in groups of exercise recommended and not recommended (i.e. SEEM < 7 or SEEM ≥ 7). ADH model provided somewhat better discriminability for ADH than that from AD model only in patients for whom exercise is not recommended (**Supplementary Figure S8)**.

## DISCUSSION

### Main findings

To the best of our knowledge, this is the first study to systematically and comprehensively evaluate the interactions of medication and other factors with the effects of exercise training in patients with chronic heart failure. The HF-ACTION trial showed no statistically significant change in the rate of AD between the exercise and usual care group.^**9**^ However, the present study’s findings suggest that exercise training had been associated with a significant and notable increase in the risk of AD in patients taking RASIs without BBs at baseline. Although exercise reportedly reduced the event rate of ADH in the entire study population in the HF-ACTION trial, this study showed that this effect was probably limited to patients taking BBs. Thus, it was demonstrated that the risk-benefit balance of exercise training varies greatly according to patient status, and a criterion is necessary to determine whether exercise training should be performed. This analysis is post-hoc in design and its limitations require consideration, as described later, it is yet important to make the best effort to improve patient outcomes from the current knowledge. With this in mind, here we proposed the SEEM score.

### Exercise effect and medications

In patients with chronic heart failure, exercise training improves functional capacity, increases muscle strength, and improves quality of life,^**1–5,21–23**^ but it also increases cardiac workload by increasing heart rate via sympathetic nervous system activation.^**14,24**^ Increasing the intensity may induce arrhythmia.^**25**^ BBs suppress sympathetic nervous system activation and reduce the increased demand for myocardial energy due to increased heart rate.^**16**^ A previous model-based meta-analysis showed that this cardiac load reduction was associated with improved prognosis of patients with chronic heart failure.^**13**^ In addition, BBs inhibit the onset of ventricular arrhythmias and prevent sudden death.^**26–28**^ Although the mechanism of the larger effect of exercise on ADH in patients taking only BBs at baseline in this study is unknown, our data likely indicate that BBs may play an important role in maximizing the benefits of exercise training, such as by improving exercise tolerance while reducing the extra load on the heart and decreasing the likelihood of arrhythmia that exercise may cause.

In contrast, our data showed that RASIs might have a negative effect on exercise. It is difficult to explain the mechanism of this effect if exists. Taking RASIs is generally considered to reduce cardiac load, but it also may reduce cardiac load at rest while increasing cardiac load during exercise.^**29**^ Volpe et al. reported that left ventricular dynamic adaptations to stress of acute volume loading are compromised for in patients with idiopathic dilated cardiomyopathy and that the impaired responses were ameliorated by enalapril treatment.^**30**^ Although the stress investigated in Volpe’s study is a volumetric rather than exercise load, such improved responses to stress by RASI might explain the increase in cardiac activity and load potentially induced by exercise.

Owing to the limited sample size, whether the effect of exercise training was similar between ACEIs and ARBs could not be statistically examined in this study. In both patient groups taking ACEIs or ARBs, it should be emphasized that there was a consistent tendency toward a reduced AD in the usual care group (**Supplementary Figures S9 and S10**). On the other hand, since the HF-ACTION trial was conducted in 2003–2008, the medication algorithms for chronic heart failure differed from the current ones. Sodium-glucose cotransporter-2 inhibitors and angiotensin receptor-neprilysin inhibitors (ARNIs) have since been demonstrated^**31**^ to improve chronic heart failure prognosis.^**7,32**^ Although the impact of exercise training on these therapies could not be evaluated in this study, given the similarity in the mechanism of action between RASIs and ARNIs, similar caution might be required when ARNIs are taken without BBs.

Current chronic heart failure guidelines recommend the combined use of RASIs and BBs as first-line therapy.^**7,32**^ On the other hand, our analysis implied that the risk in patients taking RASIs alone was the lowest for AD in the usual care group. However, the HF-ACTION trial was randomized for exercise but not for medications. Therefore, it was inappropriate to determine the superiority or inferiority of medications rather than exercise effects in the present analysis, and it will be necessary to confirm this in the future.

### Factors influencing outcomes

In addition to the interactions with exercise, many prognostic factors have been reported for chronic heart failure. O’Connor et al. reported that exercise duration in the CPX test, BUN, and sex were important for AD and ADH based on the analysis of patient information of the HF-ACTION trial.^**33**^ While their study analyzed 48 candidate factors, this study analyzed a larger number of candidate factors and selected estimated glomerular filtration rate commonly for AD and ADH, exercise duration in the CPX test for ADH, and sex for AD (**Supplementary Tables S3 and S4**). In both studies, exercise capacity, renal function, and sex were selected. Overall, although this study is novel in that it includes many interactions with exercise, the analysis of other prognostic factors was considered consistent with that of previous studies.

### Exercise recommendation based on SEEM score

The SEEM score was constructed based on the results of Cox proportional hazards analysis for AD and successfully identified patients who should and should not perform exercise training in terms of AD and ADH. The score consists of pulse pressure, history of stroke, hemoglobin concentration, abnormal electrocardiography findings, and BMI in addition to BB medications.

A pulse pressure of 30 mmHg or more, calculated by subtracting diastolic pressure from systolic pressure, was identified as the most important beneficial factor on exercise effect other than medications for both AD and ADH. In patients with advanced chronic heart failure with reduced cardiac contractility, systolic blood pressure is low;^**34**^ thus, pulse pressure is low. It is plausible that exercise causes cardiac overload and increases the risk of AD in these patients. Anemia (i.e., low hemoglobin level) is considered a prognostic factor for chronic heart failure, and in the HF-ACTION trial, it was reported that patients with anemia were at higher risk for both AD and ADH.^**35–38**^ In patients with chronic heart failure complicated by anemia, the oxygen supply is reduced by the latter.^**39**^ Thus, the prognosis of these patients may be worsened by exercise because of the additional load on the heart. Therefore, in addition to pulse pressure, the results of this analysis, which suggest that patients with low hemoglobin levels are not recommended to perform exercise training, are reasonable.

As for the ECG findings, the result that exercise is not recommended in patients with abnormal left ventricular conduction, while exercise therapy is recommended in those with normal left ventricular conduction, seems reasonable considering the risk of sudden death due to arrhythmia.^**25**^ On the other hand, the result that exercise is recommended in patients with a history of stroke is currently difficult to adequately explain. A history of stroke was a factor that increased the risk of AD but had a positive effect on exercise. Patients with a history of stroke were more likely to be prescribed antithrombotic drugs.^**40–43**^ Therefore, it is conceivable that a history of stroke, such as in patients using antithrombotic drugs, could be a confounding factor. In this study, the risk of AD was decreased by exercise training in patients with a BMI of < 25. Maintaining one’s muscle strength through exercise may contribute to the prevention of cachexia or inhibition of its progression.^**44–47**^

We considered the possibility of calculating an appropriate score from the factors selected in the Cox proportional hazards analysis for ADH rather than AD. Some of the factors that affected the endpoints of AD and ADH were common to both the main effect and the interaction, suggesting that the use of a score derived from AD or ADH could improve both in the clinical settings. When comparing HRs between patients who decided to perform exercise training according to the SEEM score and those who did not, there was a significant difference in both AD and ADH (**Figure 7**), indicating the potential clinical utility of this score.

### Limitations

Since this is a retrospective study, future prospective evaluations are needed to fully confirm its results. As mentioned above, the HF-ACTION trial was not stratified by medications; therefore, it is inappropriate to conclude the superiority or inferiority of medications together with exercise. The results of all subgroup analyses that were not randomly assigned should be considered hypothetical. In addition, the sensitivity of some medication groups was insufficient because of the limited number of patients. In particular, we integrated ACEIs and ARBs, and patients who did not use BBs or RASIs could not be analyzed.

The population included in this analysis consisted of patients with stable chronic heart failure, a reduced left ventricular ejection fraction (≤35%), NYHA class II–IV despite treatment, and the ability to perform exercise. While we believe that it is appropriate to apply the SEEM score to these patients, it may not be applicable to other populations, such as those who had a recent cardiovascular event or comorbidities or limitations that could interfere with exercise training. In addition, the appropriateness of exercise training may vary greatly by exercise type and intensity. In particular, if exercise is expected to be harmful, future studies may need to examine the consequences of reducing exercise intensity.

## CONCLUSIONS

The benefit of exercise training demonstrated in the original HF-ACTION trial was, in this study, confirmed only in a limited patient population. The choice of exercise training in patients with chronic heart failure requires careful judgment that considers patient condition and medications, as reflected in the SEEM criteria proposed in this study. Further prospective validation would be required of its usefulness in the clinical setting.

## Data Availability

The data used in this study are available from National Heart, Lung, and Blood Institute with determined procedure and permission of National Heart, Lung, and Blood Institute.

## ABBREVIATIONS

AD: all-cause death
ADH: all-cause death and hospitalization
BB: beta-blocker
CPX: clinical performance examination
RASI: angiotensin-converting enzyme inhibitor and/or angiotensin II receptor blocker
SEEM: Score for Eligibility for Exercise on Mortality.

## DECLARATIONS

### Ethics approval and consent to participate

We analyzed anonymized individual patient-level data from the HF-ACTION trial obtained from the National Heart, Lung, and Blood Institute, USA. The study protocol and design were reviewed and approved by the ethical review board of the Graduate School of Pharmaceutical Sciences at Chiba University.

### Consent for publication

Not applicable

### Competing interests

YS and SG are employees of Sanofi K.K. and Astellas Pharma Inc., respectively; however, Sanofi K.K. and Astellas Pharma Inc. were not involved in this analysis. All other authors have no conflict of interest to disclose.

### Funding

This work was supported by the AMED under Grant number JP21mk0101159.

### Authors’ contributions

Y.S. and A.H. designed research; Y.S., H.Y. and, S.G. analyzed data; A.H., Y.S., Y.F. and N.A. conceived and oversaw the study; and Y.S., H.Y., H.S., H.H., and A.H. wrote the paper.

## Acknowledgements

This manuscript was prepared using HF-ACTION Research Materials obtained from the BioLINCC data base of NHLBI Biologic Specimen and Data Repository Information Coordinating Center (Bethesda, MD, USA) and does not necessarily reflect the opinions or views of the HF-ACTION or the NHLBI.

## ADDITIONAL FILE

Additional file 1: Supplemental Materials (Supplementary Statistical Methods, Supplementary Tables S1–S6, Supplementary Figures S1–S10)

## Notes

### Author Declarations

The study protocol and design were reviewed and approved by the ethical review board of the Graduate School of Pharmaceutical Sciences at Chiba University.

