## Supplemental Materials for "Exercise Training Outcomes in Patients with Chronic Heart Failure with Reduced Ejection Fraction Depend on Medications and Disease Conditions: Proposal of SEEM Score to Judge Exercise Suitability"

### Supplementary Statistical Methods

#### Model Development

The Cox models in this analysis have the following basic structure:

$$\lambda\left( t \right)=\lambda_{0}\left( t \right)\exp\left( \theta\right)$$

$$\begin{aligned} \theta=(&\beta_{1}*exercise*rasi1\_beta1+ \\ &\beta_{2}*exercise*rasi1\_beta0+ \\ &\beta_{3}*exercise*rasi0\_beta1+ \\ &\beta_{4}*exercise*rasi0\_beta0+ \\ &\beta_{5}*rasi1\_beta1+ \\ &\beta_{6}*rasi1\_beta0+ \\ &\beta_{7}*rasi0\_beta1+ \\ &\beta_{8}*rasi0\_beta0) \end{aligned}$$

where, the variables are defined as follows:

| exercise | 1 if in the exercise training group, 0 otherwise |
| --- | --- |
| rasi1_beta1 | 1 if treated with both RASI and BB at baseline, 0 otherwise |
| rasi1_beta0 | 1 if treated with RASI but not BB at baseline, 0 otherwise |
| rasi0_beta1 | 1 if treated with BB but not RASI at baseline, 0 otherwise |
| rasi0_beta0 | 1 if treated with neither RASI nor BB at baseline, 0 otherwise |

Since the number of patients who were not taking both RASI and BB at baseline was extremely small (n=10; 0.5%), the exercise effect in this group (β4) was fixed at 0. The coefficient for "rasi1_beta1" (β5) was also fixed at 0 as this group was handled as a tentative reference.

Factors affecting exercise effects other than medication at baseline were comprehensively explored. Candidate variables are listed in **Supplementary Table S1**. First, to screen possible covariate effects, simple Cox models were developed including treatment group (i.e., dummy variable indicating exercise training or usual care), variable of interest (i.e., one of the candidate variables listed in Supplementary Table S1), and their interaction terms. If the interaction was significant at a P value less than 0.3, both the interaction term and the main effect were entered into the subsequent covariate selection process. If the P value for interaction was greater than 0.3 but the main effect of the variable achieved a P value of less than 0.3, only the main effect was entered into the subsequent process. The full model was developed by applying the forward inclusion method to the selected candidates of interaction effects and main effects at a significance level of P=0.1. The backward elimination method with a significance level of P=0.05 was then applied to the full model to obtain the final model. To ensure the robustness of the model, the backward elimination was repeated using 1,000 bootstrap datasets. The main effect was considered eligible for inclusion in the final model if it was adopted in more than 800 times (>80%) of the bootstrap replications, while more than 600 times (>60%) for the interaction effect. To avoid computational divergence due to population imbalance, the bootstrapping was performed after stratifying data by treatment group, medication group, and death or hospitalization at 4 years.

The methods described above imply multiple interaction terms between exercise and patient characteristics will be incorporated into the same single model. In such a model, the main effect of exercise represents the effects in a very small population that is not applicable to any of the interaction factors included in the model; in other words, the estimate of the exercise effect becomes meaningless as the number of interaction factors increases. To address this issue, the Cox regression analysis was performed after all the variables other than the treatment group and medication groups were zero-centered (i.e., subtracting the mean of each variable). This adjustment does not change the coefficient estimates and significance of the main effects or interactions for these variables. However, the main effect of exercise training will be estimated as values in a hypothetical average population with the means of all interaction factors. For example, suppose a Cox model that includes the interaction of age (continuous variable) and exercise (binary variable). Usually, the estimated coefficient for the main effect of exercise represents the effect of exercise when the age is zero. In contrast, if the variable for age is corrected in advance so that the average value is zero, the estimated main effect of exercise will represent the effect of exercise in a population with an average age.

### Supplementary Figures

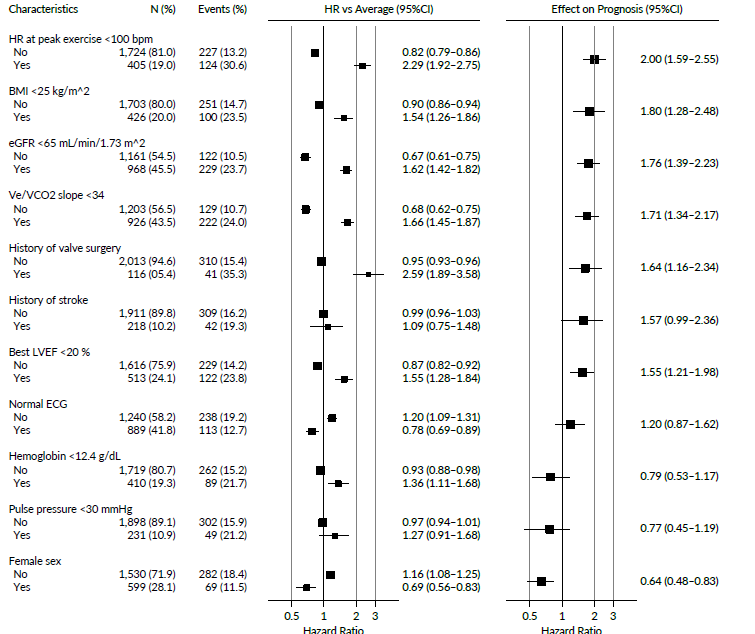

#### Figure S1. Influence of Main Effect Other than Interaction with Exercise for All-Cause Death

All the HRs, effects and their CIs were calculated from the analysis of 1,000 bootstrap datasets based on the multivariate Cox proportional hazards model analysis. BMI Body mass index; CI Confidence interval; ECG Electrocardiogram; eGFR estimated glomerular filtration rate; HR Hazard ratio; HR Heart rate; LVEF Left ventricle ejection fraction; Ve/VCO2 Ventilation/carbon dioxide production ratio.

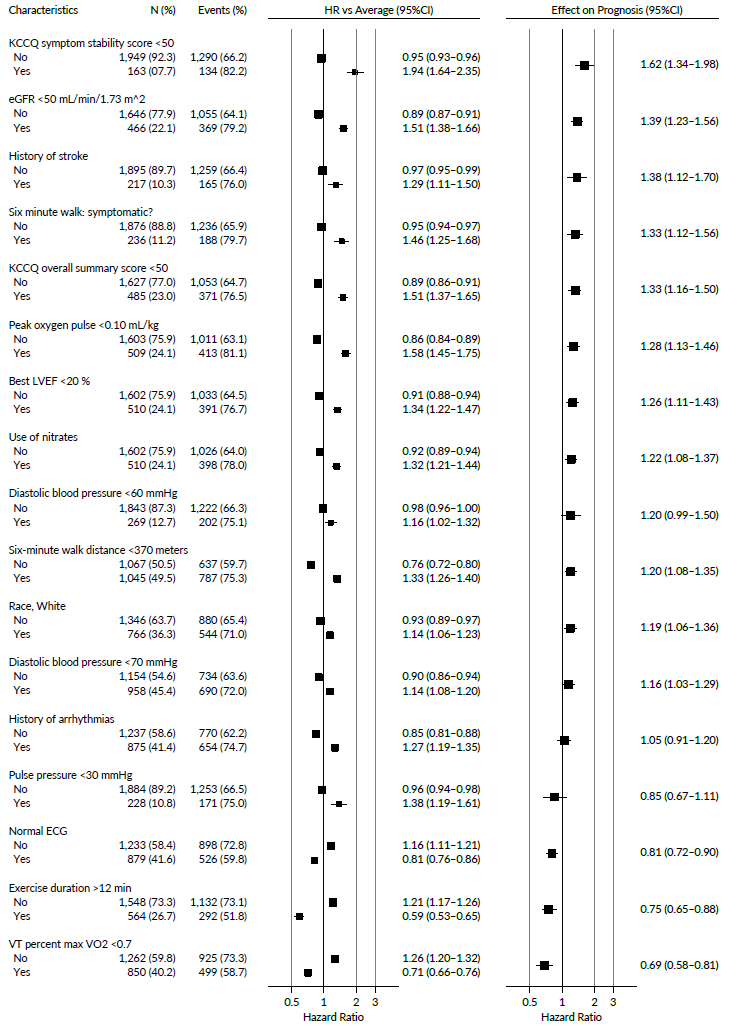

#### Figure S2. Influence of Main Effect Other than Interaction with Exercise for All-Cause Death or All-Cause Hospitalization

All the HRs, effects and their CIs were calculated from the analysis of 1,000 bootstrap datasets based on the multivariate Cox proportional hazards model analysis. CI Confidence interval; ECG Electrocardiogram; eGFR Estimated glomerular filtration rate; HR Hazard ratio; KCCQ Kansas City Cardiomyopathy Questionnaire; LVEF Left ventricle ejection fraction; VT Ventilatory threshold.

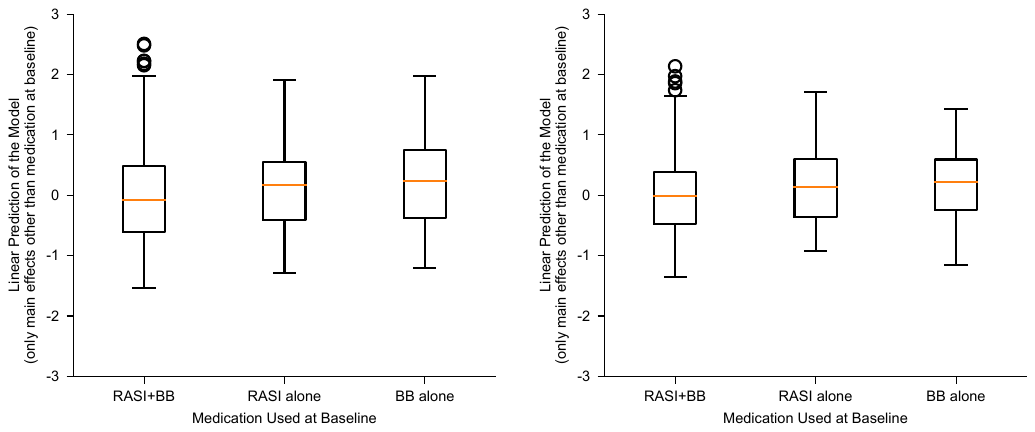

#### Figure S3. Distributions of Linear Predictors Calculated Using Main Effects Other than Baseline Medication

Left: all-cause death, right: all-cause death or all-cause hospitalization. The box represents the range from the first quartile (Q1) to the third quartile (Q3) of the data. The whiskers extend from the box by 1.5x the inter-quartile range. Flier points are those past the end of the whiskers. BBs, beta-blockers; RASIs, angiotensin-converting enzyme inhibitors and/or angiotensin II receptor blockers.

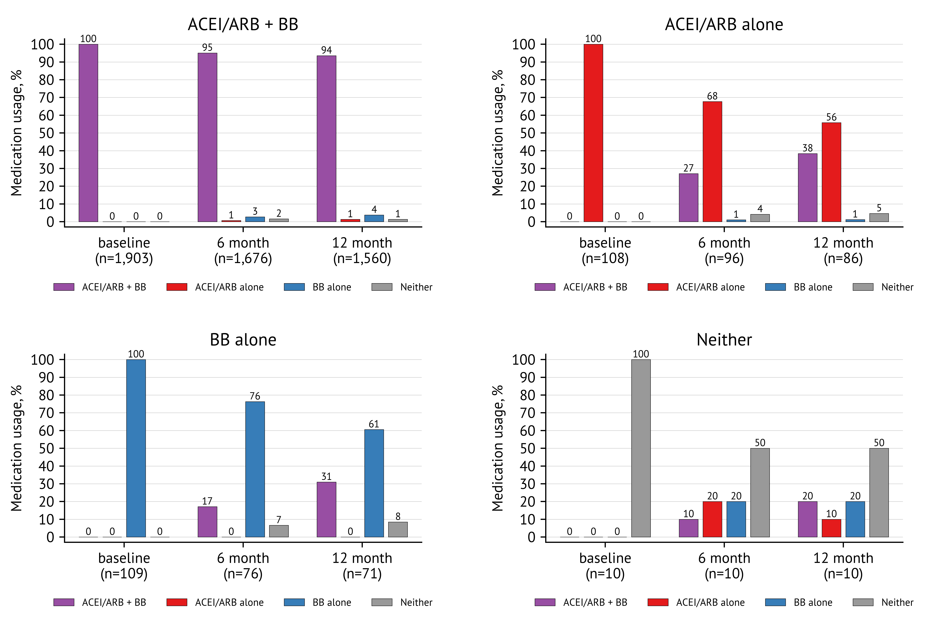

#### Figure S4. Percentage Change in Medication Category from Baseline

Percentage change in medication category from baseline at 3, 6, and 12 months after randomization are shown. A: Patients using both RASI and BB at baseline, B: Patients using RASI alone at baseline, C: Patients using BB alone at baseline, D: Patients using neither RASI nor BB at baseline. BB Beta-blocker; RASI Angiotensin-converting enzyme inhibitor and/or angiotensin II receptor blocker.

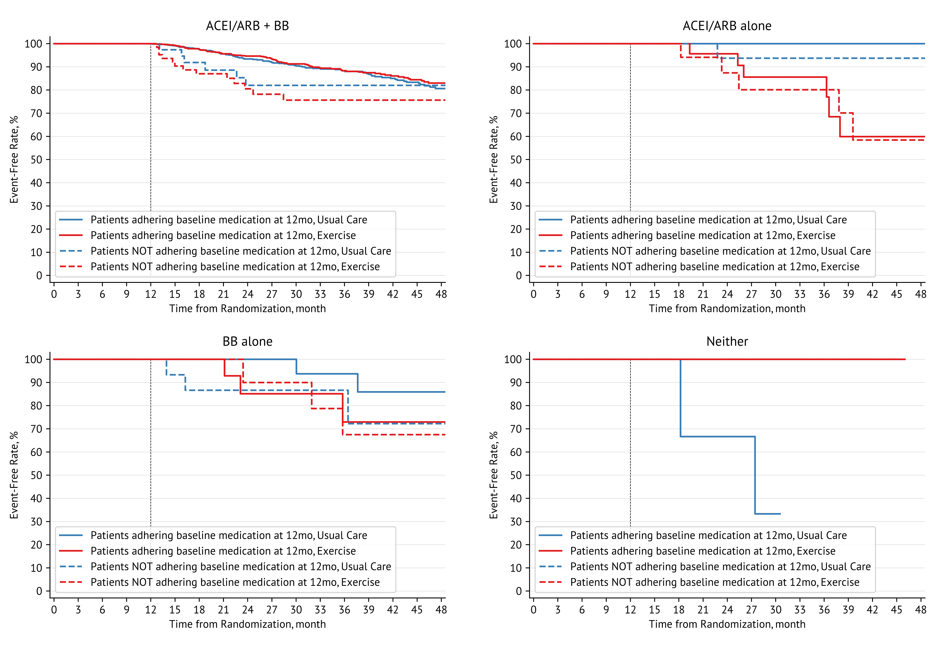

#### Figure S5. Kaplan-Meier Curves for All-Cause Death in patients adhering or not adhering baseline medication

Survival curves for exercise effects in patients adhering or not adhering baseline medication after 12 months from randomization are shown. A: Patients using both RASI and BB at baseline, B: Patients using RASI alone at baseline, C: Patients using BB alone at baseline, D: Patients using neither RASI nor BB at baseline. BB Beta-blocker; RASI Angiotensin-converting enzyme inhibitor and/or angiotensin II receptor blocker.

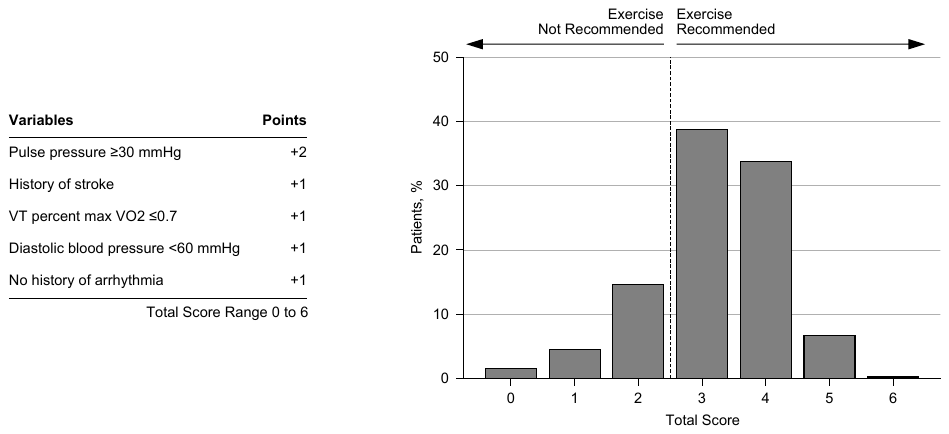

#### Figure S6. Elements of Score Calculated from Endpoint of All-Cause Death or All-Cause Hospitalization and Distribution of Score Among HF-ACTION Study Patients

The scores incorporated all factors included in the interaction terms with exercise in the final model of Cox proportional hazards model, as well as medication groups for which the exercise effect was estimated to be statistically significantly different from that in the overall study population. For ease of use, all variables were assigned integer points by scaling the β coefficients while maintaining the gradient of the effects. VT Ventilatory threshold; VO2 Oxygen uptake.

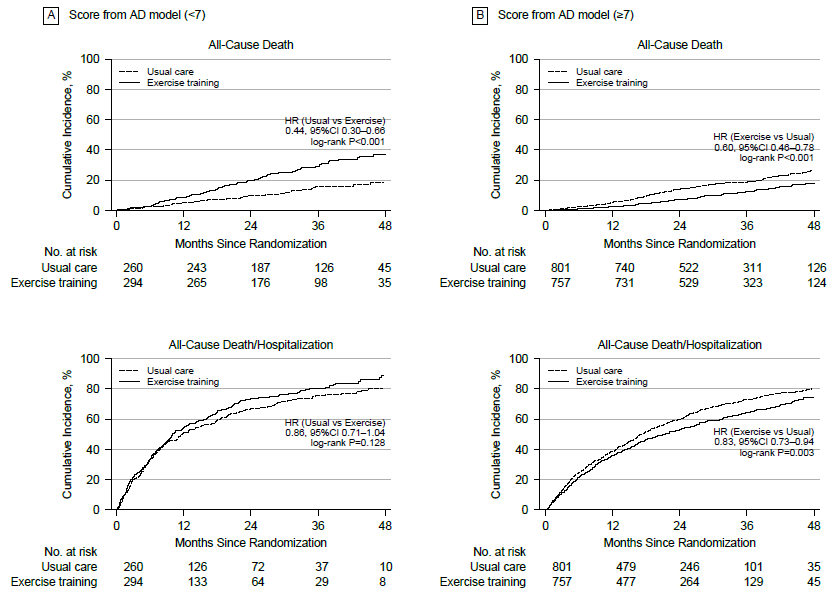

#### Figure S7. Cumulative incidence for each endpoint by lower and upper SEEM score group

Cumulative incidence for each endpoint by lower and upper SEEM score group was calculated. A: Lower score group (<7), B: Upper score group (≥7). CI Confidence interval; HR Hazard ratio.

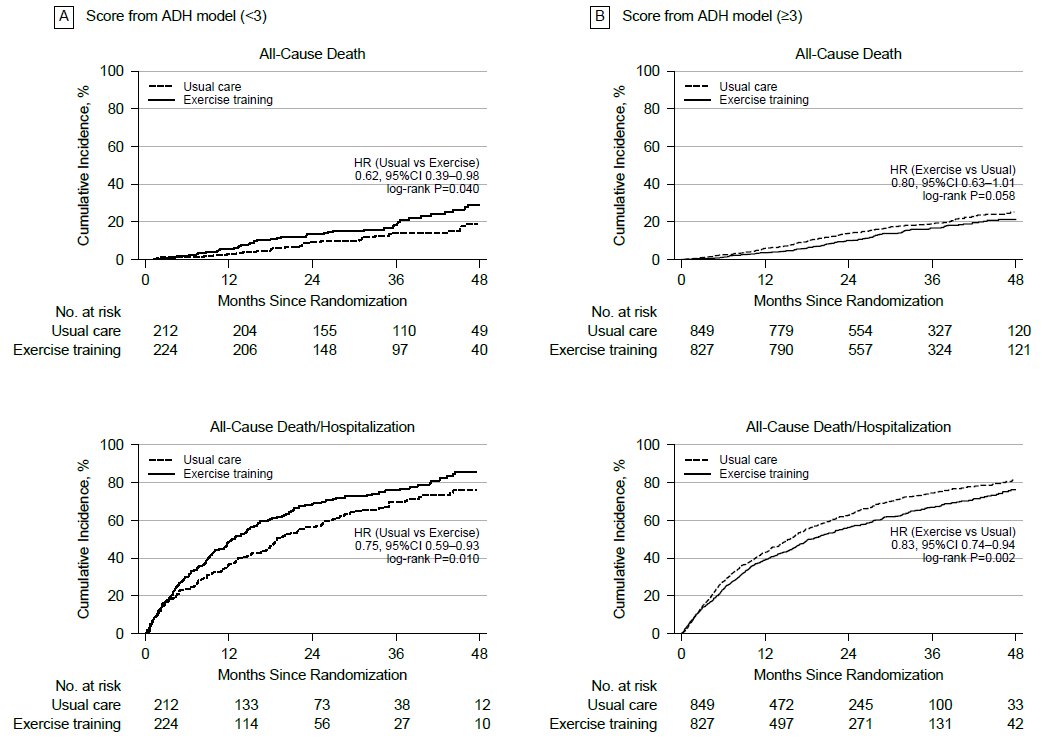

#### Figure S8. Cumulative incidence for each endpoint by lower and upper score group developed from ADH model

Cumulative incidence for each endpoint by lower and upper score group was calculated by ADH model. A: Lower score group (<3), B: Upper score group (≥3). CI Confidence interval; HR Hazard ratio.

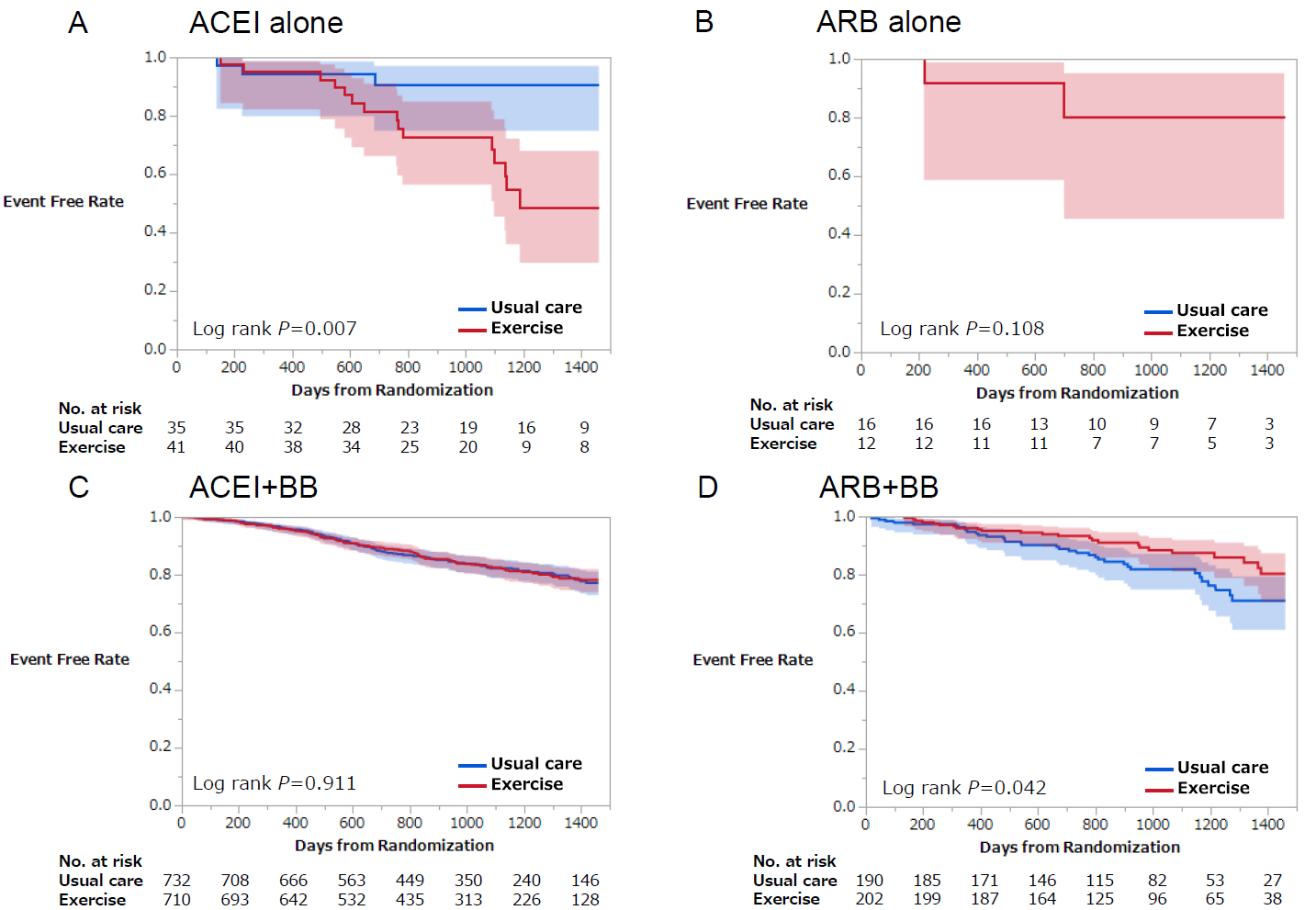

#### Figure S9. Kaplan–Meier Curve for All-Cause Death by Medication at Baseline (Separating ACEI and ARB)

Kaplan-Meier curves of each medication group for all-cause death was directly compared. A: Patients using ACEI alone at baseline, B: Patients using ARB alone at baseline, C: Patients using both ACEI and BB at baseline, D: Patients using both ARB and BB at baseline. Shaded area represents 95% confidence intervals. ACEI Angiotensin-converting enzyme inhibitor; ARB Angiotensin II receptor blocker; BB Beta- blocker.

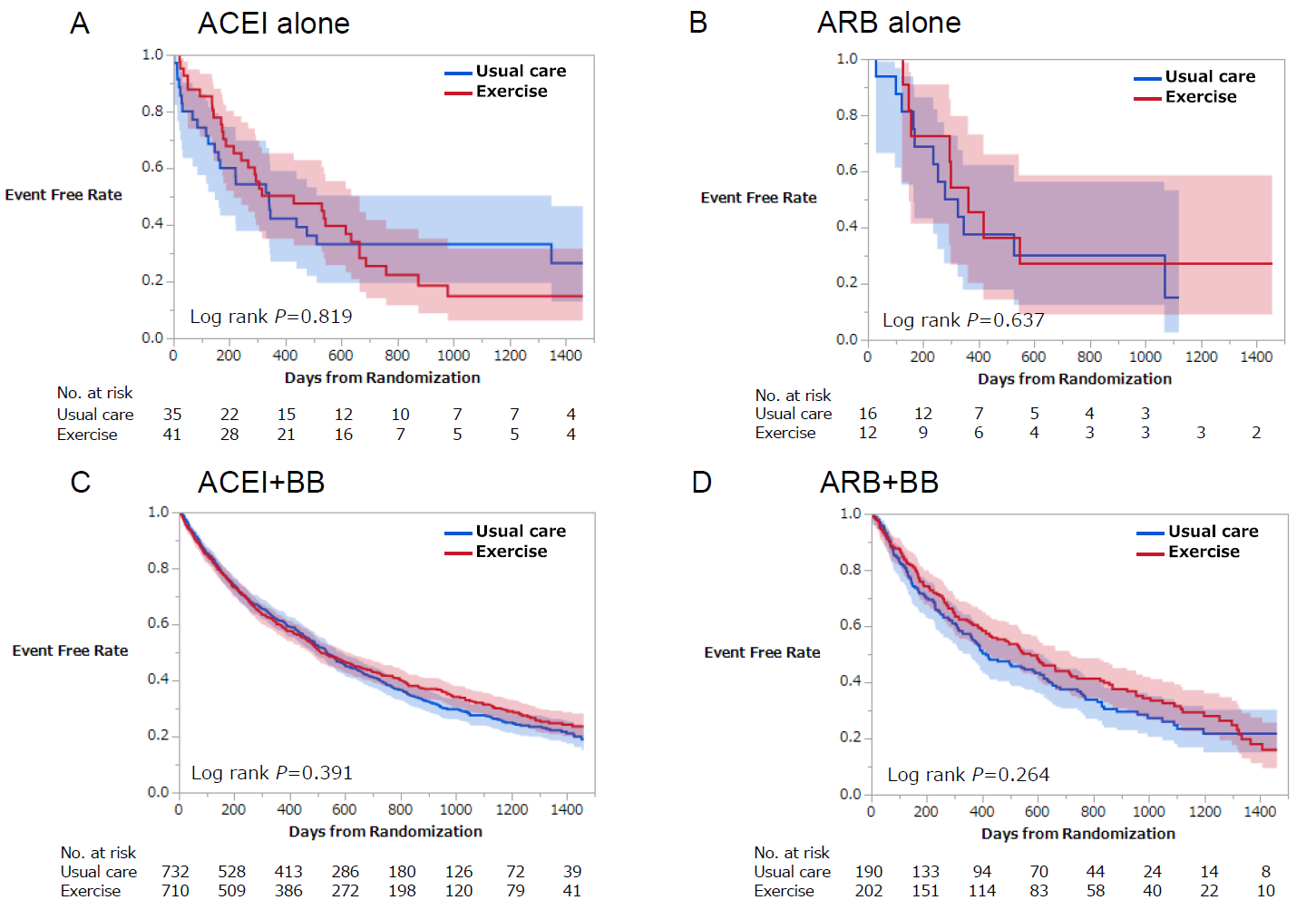

#### Figure S10. Kaplan–Meier Curve for All-Cause Death and All-Cause Hospitalization by Medication at Baseline (Separating ACEI and ARB)

Kaplan-Meier curves of each medication group for all-cause death or all-cause hospitalization was directly compared. A: Patients using ACEI alone at baseline, B: Patients using ARB alone at baseline, C: Patients using both ACEI and BB at baseline, D: Patients using both ARB and BB at baseline. Shaded area represents 95% confidence intervals. ACEI Angiotensin-converting enzyme inhibitor; ARB Angiotensin II receptor blocker; BB Beta-blocker.

### Supplementary Tables

#### Table S1. List of Variables Used as Candidate Covariates in Cox Regression Analysis

| **Domain** | **No** | **Variable** | **Label** | **Type** | **Cutoff** | **Unit** | **Missing (%)** | **Excluded** |
| --- | --- | --- | --- | --- | --- | --- | --- | --- |
| **Demographics** | **1** | **age** | Age | continuous | 50;60;70 | years | 0 |  |
|  | **2** | **male** | Mele sex | categorical |  |  | 0 |  |
|  | **3** | **white** | Race, White | categorical |  |  | 1.5 |  |
|  | **–** | **black** | Race, Black | categorical |  |  | 1.5 | Correlated with white (chi square >300) |
|  | **4** | **asian** | Race, Asian, Amer Ind, Pac. Isl | categorical |  |  | 1.5 |  |
| **Vital Sign** | **5** | **bmi** | Body mass index | continuous | 25;30;35 | kg/m^2 | 0.2 |  |
|  | **–** | **weightkg** | Body weight | continuous | 75;90;110 | kg | 0 | Correlated with bmi (spearman >0.8) |
|  | **6** | **basehr** | Heart rate | continuous | 60;70;80 | bpm | 0.2 |  |
|  | **7** | **pulsepr** | Pulse pressure | continuous | 30;40;50 | mmHg | 0.2 |  |
|  | **8** | **sbp** | Systolic blood pressure | continuous | 100;110;125 | mmHg | 0.2 |  |
|  | **9** | **dbp** | Diastolic blood pressure | continuous | 60;70;80 | mmHg | 0.2 |  |
|  | **–** | **egfr** | eGFR | continuous | 50;65;80 | mL/min/1.73 m^2 | 10.2 | Correlated with creatnc (spearman >0.8) |
| **Subject Status** | **10** | **bestlvef** | Best available LVEF | continuous | 20;25;30 | % | 0.2 |  |
|  | **11** | **nyhacl** | NHYA class | continuous | 3;4 |  | 0 |  |
|  | **12** | **beckb** | Beck score | continuous | 4;8;15 |  | 0.4 |  |
|  | **13** | **ccsangcl** | Current CCS angina class | continuous | 1;2 |  | 0.1 |  |
|  | **14** | **hfhosp** | No. of HF hospitalizations in last 6 months | continuous | 1;2;3 |  | 0.8 |  |
|  | **15** | **hospsxmo** | No. of hospitalizations in last 6 months | continuous | 1;2;3 |  | 0.7 |  |
|  | **–** | **ischemic** | Ischemic etiology | categorical |  |  | 0 | Correlated with llwa (chi square >300) |
|  | **16** | **renaldys** | Renal dysfunction | categorical |  |  | 10.2 |  |
|  | **17** | **smokenever** | Never smoker | categorical |  |  | 0.5 |  |
|  | **18** | **smokecurr** | Current smoker | categorical |  |  | 0.5 |  |
|  | **–** | **alcohol** | Alcohol use | categorical |  |  | 1.8 | Correlated with nodrink (chi square >300) |
|  | **19** | **nodrink** | Patient no longer drinks | categorical |  |  | 0 |  |
| **ECG** | **20** | **ecgcond3** | Normal ECG | categorical |  |  | 2.5 |  |
| **CPX** | **21** | **cpxdur** | Exercise duration in CPX test | continuous | 7;10;12 | minutes | 0.9 |  |
|  | **22** | **peakvo2** | Peak VO2 | continuous | 11;14;17 | mL/kg/min | 2.4 |  |
|  | **23** | **vevco2** | Ve/VCO2 slope | continuous | 24;30;34 |  | 3.2 |  |
|  | **24** | **peakrer** | Peak respiratory exchange ratio | continuous | 1.0;1.1;1.2 |  | 3.1 |  |
|  | **–** | **vo2atvt** | VO2 at ventilatory threshold | continuous | 8.8;10.6;12.5 | mL/min/kg | 16.8 | Correlated with peakvo2 (spearman >0.8) |
| **CPX Test** | **25** | **cpxrsthr** | Resting heart rate | continuous | 60;70;80 | bpm | 0.1 |  |
|  | **26** | **hrpeakex** | HR at peak exercise | continuous | 100;120;130 | bpm | 0.1 |  |
|  | **–** | **cpxhrr** | Heart rate reserve | continuous | 35;50;60 | bpm | 0.1 | Correlated with hrpeakex (spearman >0.8) |
|  | **27** | **peakrpe** | Borg RPE score at peak exercise | continuous | 15;17;19 |  | 0.5 |  |
|  | **28** | **vtpctvo2** | VT percent max VO2 | continuous | 0.6;0.7;0.8 |  | 16.8 |  |
|  | **29** | **pko2puls** | Peak oxygen pulse | continuous | 0.10;0.12;0.15 | mL/kg | 2.4 |  |
|  | **–** | **tmrerone** | Time to RER = 1.0 | continuous | 6;8;10 | minutes | 22.7 | Correlated with cpxdur (spearman >0.8) |
|  | **30** | **hrwl2** | Heart rate at the end of WL 2 | continuous | 80;90;100 | bpm | 5.3 |  |
|  | **–** | **ouesall** | Oxygen uptake efficiency slope measured from entire CPX test | continuous | 1200;1600;2000 | mL/min/log(L/min) | 2.6 | Correlated with abspkvo2 (spearman >0.8) |
|  | **–** | **ouesupto6** | Oxygen uptake efficiency slope measured from the first 6 minutes of CPX test | continuous | 1200;1600;2000 | mL/min/log(L/min) | 18.5 | Correlated with ouesall (spearman >0.8) |
|  | **–** | **ouesuptorer1** | Oxygen uptake efficiency slope measured during CPX test while RER = 1.0 | continuous | 1200;1600;2000 | mL/min/log(L/min) | 27 | Correlated with ouesall (spearman >0.8) |
|  | **–** | **weber** | Weber class | continuous | 2;3;4 |  | 2.4 | Correlated with peakvo2 (spearman >0.8) |
|  | **31** | **abspkvo2** | Absolute peak VO2 | continuous | 1000;1300;1700 | mL/min | 2.5 |  |
| **6-Min Walk Test** | **32** | **sixmwlka** | Six-minute walk: able to walk?, Yes | categorical |  |  | 0 |  |
|  | **33** | **sixmwlks** | Six minute walk: symptomatic?, Yes | categorical |  |  | 3.8 |  |
|  | **34** | **sixmwlkd** | Six-minute walk distance, subjects able to walk | continuous | 300;370;440 | meters | 2.2 |  |
|  | **35** | **walkwork** | Work done during 6MW, subjects able to walk | continuous | 240;320;400 | kJ | 2.2 |  |
| **Echocardiography** | **–** | **eprimevel** | E prime velocity | continuous |  |  | 60.6 | Missing in >30% of samples |
|  | **–** | **eeprimevelrat** | E/E prime velocity ratio | continuous |  |  | 65.4 | Missing in >30% of samples |
|  | **–** | **ewavsept** | Tissue Doppler E wave (septal annulus) | continuous |  |  | 51.1 | Missing in >30% of samples |
|  | **–** | **ewavlat** | Tissue Doppler E wave (lateral annulus) | continuous |  |  | 50 | Missing in >30% of samples |
|  | **–** | **mvpkevel** | Doppler MV peak E velocity | continuous |  |  | 33.2 | Missing in >30% of samples |
|  | **–** | **lveddt** | Left ventricular early diastolic deceleration time | continuous |  |  | 30.8 | Missing in >30% of samples |
|  | **–** | **lftatrdm** | Left atrial dimension | continuous | 4.0;4.5;5.0 | cm | 29.2 |  |
|  | **37** | **lftvmass** | Left ventricular mass | continuous | 200;300;400 | g | 29.3 |  |
|  | **38** | **mitregrg** | Mitral regurgitation | continuous | 10;20;30 | % | 22.3 |  |
|  | **39** | **lftvdiad** | Left ventricular diastolic dimension | continuous | 5.9;6.6;7.4 | cm | 29.4 |  |
|  | **–** | **eavelrat** | MV peak E velocity / A velocity | continuous |  |  | 33.1 | Missing in >30% of samples |
|  | **–** | **mvpkevlv** | MV peak E velocity - Valsalva | continuous |  |  | 75.1 | Missing in >30% of samples |
|  | **–** | **mvpkavlv** | MV peak A velocity - Valsalva | continuous |  |  | 75.1 | Missing in >30% of samples |
|  | **–** | **mvearatv** | MV E/A ratio - Valsalva | continuous |  |  | 75 | Missing in >30% of samples |
| **Laboratory Value** | **40** | **creatnc** | Creatinine | continuous | 1.0;1.2;1.5 | mg/dL | 10.2 |  |
|  | **41** | **bun** | BUN | continuous | 15;20;30 | mg/dL | 13 |  |
|  | **–** | **totcholc** | Total cholesterol | continuous |  | mg/dL | 35.4 | Missing in >30% of samples |
|  | **–** | **hdl** | High-density lipoprotein | continuous |  | mg/dL | 38.5 | Missing in >30% of samples |
|  | **–** | **ldl** | Low-density lipoprotein | continuous |  | mg/dL | 39.9 | Missing in >30% of samples |
|  | **–** | **hema1cc** | Hemoglobin A1c | continuous |  | % | 73.4 | Missing in >30% of samples |
|  | **–** | **bnp** | BNP | continuous |  | pg/mL | 64.2 | Missing in >30% of samples |
|  | **–** | **probnp** | Pro-BNP | continuous |  | pg/mL | 96.5 | Missing in >30% of samples |
|  | **42** | **hemogloc** | Hemoglobin | continuous | 12.4;13.5;14.6 | g/dL | 24.1 |  |
| **Medical History** | **43** | **valvsurg** | History of prior valve surgery | categorical |  |  | 0 |  |
|  | **44** | **revasc** | History of prior revascularization | categorical |  |  | 0 |  |
|  | **–** | **priorpci** | History of prior PCI | categorical |  |  | 0 | Correlated with ischemic (chi square >300) |
|  | **–** | **cabg** | History of prior CABG | categorical |  |  | 0 | Correlated with ischemic (chi square >300) |
|  | **45** | **copd** | History of COPD | categorical |  |  | 0.9 |  |
|  | **–** | **diabetes** | History of diabetes | categorical |  |  | 0 | Correlated with insulin (chi square >300) |
|  | **–** | **depress** | History of depression | categorical |  |  | 0 | Correlated with ssri (chi square >300) |
|  | **46** | **angina** | History of angina | categorical |  |  | 0 |  |
|  | **–** | **priormi** | History of myocardial infarction | categorical |  |  | 0 | Correlated with ischemic (chi square >300) |
|  | **–** | **pvd** | History of peripheral vascular disease | categorical |  |  | 0.5 | Correlated with claud (chi square >300) |
|  | **47** | **stroke** | History of stroke | categorical |  |  | 0 |  |
|  | **48** | **hyperten** | History of hypertension | categorical |  |  | 0.6 |  |
|  | **–** | **arr** | History of arrhythmias | categorical |  |  | 0 | Correlated with vtach (chi square >300) |
|  | **–** | **afibflut** | History of atrial fibrillation or flutter | categorical |  |  | 0 | Correlated with arr (chi square >300) |
|  | **49** | **brady** | History of symptomatic bradycardia | categorical |  |  | 0 |  |
|  | **50** | **vtach** | History of sustained ventricular tachycardia/ventri | categorical |  |  | 0 |  |
|  | **–** | **hlipid** | History of hyperlipidemia | categorical |  |  | 0.1 | Correlated with llwa (chi square >300) |
|  | **51** | **claud** | History of claudication | categorical |  |  | 0 |  |
|  | **52** | **cancer** | History of cancer in last 5 years | categorical |  |  | 0.6 |  |
|  | **–** | **aicd** | On an automatic implantable cardioverter-defibrillator at baseline | categorical |  |  | 0 | Correlated with vtach (chi square >300) |
|  | **53** | **bivpacer** | On a bi-ventricular pacemaker at baseline | categorical |  |  | 0 |  |
|  | **54** | **pacer** | On a pacemaker at baseline | categorical |  |  | 0 |  |
| **Medication** | **55** | **loopdiur** | Use of loop diuretic at baseline | categorical |  |  | 0 |  |
|  | **56** | **nonloop** | Use of non-loop diuretic (excluding aldosterone antagonist) at baseline | categorical |  |  | 0 |  |
|  | **–** | **hmgcoari** | Use of HMG-CoA reductase inhibitor at baseline | categorical |  |  | 0 | Correlated with llwa (chi square >300) |
|  | **57** | **digoxin** | Use of digoxin at baseline | categorical |  |  | 0 |  |
|  | **58** | **nitrate** | Use of nitrates at baseline | categorical |  |  | 0 |  |
|  | **59** | **calcchbl** | Use of calcium channel blocker at baseline | categorical |  |  | 0 |  |
|  | **60** | **insulin** | Use of insulin at baseline | categorical |  |  | 0 |  |
|  | **–** | **spiro** | Use of spironolactone at baseline | categorical |  |  | 0 | Correlated with aldant (chi square >300) |
|  | **61** | **epleren** | Use of eplerenone at baseline | categorical |  |  | 0 |  |
|  | **62** | **aldant** | Use of aldosterone antagonist (spiro. or epleren.) at baseline | categorical |  |  | 0 |  |
|  | **63** | **aspirin** | Use of aspirin at baseline | categorical |  |  | 0 |  |
|  | **64** | **llwa** | Use of liquid-lowering agent at baseline | categorical |  |  | 0 |  |
|  | **65** | **arrhyt** | Use of antiarrhythmic at baseline | categorical |  |  | 0 |  |
|  | **66** | **ssri** | Use of SSRI at baseline | categorical |  |  | 0 |  |
| **Questionnaire** | **–** | **kccqosb** | KCCQ overall summary score at baseline | continuous | 50;65;80 |  | 0 | Correlated with kccqslsb (spearman >0.8) |
|  | **–** | **kccqcsb** | KCCQ clinical summary score at baseline | continuous | 60;75;90 |  | 0 | Correlated with kccqplb (spearman >0.8) |
|  | **67** | **kccqplb** | KCCQ physical limitation score at baseline | continuous | 50;70;90 |  | 0.8 |  |
|  | **68** | **kccqslsb** | KCCQ social limitation score at baseline | continuous | 40;60;90 |  | 1.7 |  |
|  | **–** | **kccqtsb** | KCCQ total symptom score at baseline | continuous | 60;75;90 |  | 0 | Correlated with kccqsfb (spearman >0.8) |
|  | **69** | **kccqssb** | KCCQ symptom stability score at baseline | continuous | 25;50;75 |  | 0.6 |  |
|  | **70** | **kccqsfb** | KCCQ symptom frequency score at baseline | continuous | 60;75;90 |  | 0.1 |  |
|  | **–** | **kccqsbb** | KCCQ symptom burden score at baseline | continuous | 60;75;90 |  | 0 | Correlated with kccqsfb (spearman >0.8) |
|  | **71** | **kccqseb** | KCCQ self-efficacy score at baseline | continuous | 70;90;100 |  | 0.1 |  |
|  | **72** | **kccqqolb** | KCCQ quality of life score at baseline | continuous | 40;60;80 |  | 0.1 |  |
|  | **73** | **eurother** | EuroQol thermometer response | continuous | 50;70;80 |  | 2.5 |  |

#### Table S2. Patient Characteristics by Use of RASI and BB at Baseline

|  | **RASI +BB** | **RASI Alone** | **BB Alone** | **Neither** |  |
| --- | --- | --- | --- | --- | --- |
|  | **N=1888**  **(89.5%)** | **N=107**  **(5.1%)** | **N=107**  **(5.1%)** | **N=10**  **(0.5%)** |  |
| **Variable** |  |  |  |  | **P** |
| Treatment group assignment |  |  |  |  |  |
| Usual Care | 946 (49.7) | 53 (49.1) | 67 (61.5) | 4 (40.0) | 0.056 |
| Exercise | 957 (50.3) | 55 (50.9) | 42 (38.5) | 6 (60.0) | 0.056 |
| Demographics |  |  |  |  |  |
| Age, year | 58 (50–67) | 64 (55–73) | 62 (50–72) | 62 (57–64) | <0.001 |
| Male sex | 1,362 (71.6) | 82 (75.9) | 81 (74.3) | 6 (60.0) | 0.527 |
| Race |  |  |  |  |  |
| White | 1,165 (61.2) | 74 (68.5) | 75 (68.8) | 8 (80.0) | 0.101 |
| Black | 609 (32.0) | 27 (25.0) | 32 (29.4) | 1 (10.0) | 0.277 |
| Other | 102 (5.4) | 4 (3.7) | 1 (0.9) | 0 (0.0) | 0.097 |
| Region |  |  |  |  |  |
| West USA | 217 (11.4) | 16 (14.8) | 14 (12.8) | 4 (40.0) | 0.518 |
| Midwest USA | 604 (31.7) | 22 (20.4) | 33 (30.3) | 3 (30.0) | <0.05 |
| Northeast USA | 202 (10.6) | 11 (10.2) | 12 (11.0) | 0 (0.0) | 0.981 |
| South USA | 647 (34.0) | 47 (43.5) | 46 (42.2) | 2 (20.0) | <0.05 |
| Canada | 163 (8.6) | 8 (7.4) | 3 (2.8) | 1 (10.0) | 0.094 |
| France | 70 (3.7) | 4 (3.7) | 1 (0.9) | 0 (0.0) | 0.315 |
| Highest level of education |  |  |  |  |  |
| Less than high school | 230 (12.1) | 18 (16.7) | 6 (5.5) | 1 (10.0) | <0.05 |
| High school graduate or equivalent (e.g., GED) | 504 (26.5) | 32 (29.6) | 36 (33.0) | 1 (10.0) | 0.266 |
| Completed some college, but no degree | 510 (26.8) | 26 (24.1) | 30 (27.5) | 3 (30.0) | 0.807 |
| Completed associate degree/diploma program | 161 (8.5) | 12 (11.1) | 10 (9.2) | 1 (10.0) | 0.621 |
| College graduate (e.g., B.A., B.S.) | 294 (15.4) | 15 (13.9) | 13 (11.9) | 3 (30.0) | 0.565 |
| Completed graduate school (e.g, M.S., M.D., Ph.D.) | 164 (8.6) | 4 (3.7) | 8 (7.3) | 1 (10.0) | 0.184 |
| Most recent annual pre-tax household income |  |  |  |  |  |
| Less than $15,000 | 375 (19.7) | 21 (19.4) | 24 (22.0) | 3 (30.0) | 0.837 |
| $15,000–$24,999 | 307 (16.1) | 27 (25.0) | 19 (17.4) | 0 (0.0) | 0.054 |
| $25,000–$34,999 | 256 (13.5) | 11 (10.2) | 16 (14.7) | 0 (0.0) | 0.572 |
| $35,000–$44,999 | 254 (13.3) | 13 (12.0) | 18 (16.5) | 1 (10.0) | 0.582 |
| $45,000–$59,999 | 268 (14.1) | 9 (8.3) | 10 (9.2) | 3 (30.0) | 0.093 |
| $50,000–$74,999 | 125 (6.6) | 9 (8.3) | 5 (4.6) | 0 (0.0) | 0.536 |
| $75,000–$99,999 | 120 (6.3) | 4 (3.7) | 3 (2.8) | 2 (20.0) | 0.186 |
| $100,000 or more | 0 (0.0) | 0 (0.0) | 0 (0.0) | 0 (0.0) | 1.000 |
| Decline to answer | 191 (10.0) | 14 (13.0) | 14 (12.8) | 1 (10.0) | 0.421 |
| Vital Signs |  |  |  |  |  |
| Weight, kg | 90 (76–106) | 84 (72–97) | 87 (76–102) | 84 (64–89) | 0.098 |
| Body Mass Index | 30 (26–35) | 28 (25–32) | 29 (25–34) | 27 (24–29) | 0.116 |
| Heart rate, bpm | 70 (62–76) | 72 (66–80) | 72 (66–80) | 80 (68–84) | <0.001 |
| Systolic blood pressure, mmHg | 110 (100–125) | 112 (102–128) | 112 (102–130) | 110 (102–122) | 0.261 |
| Diastolic blood pressure, mmHg | 70 (60–78) | 70 (62–80) | 70 (62–78) | 70 (64–72) | 0.831 |
| Pulse pressure, mmHg | 42 (34–50) | 42 (34–55) | 42 (34–58) | 36 (32–43) | 0.377 |
| Best available LVEF | 25 (20–30) | 24 (19–29) | 23 (19–30) | 21 (17–25) | 0.133 |
| NYHA Class |  |  |  |  |  |
| NYHA II | 1,226 (64.4) | 59 (54.6) | 61 (56.0) | 5 (50.0) | <0.05 |
| NYHA III | 661 (34.7) | 48 (44.4) | 45 (41.3) | 5 (50.0) | 0.054 |
| NYHA IV | 16 (0.8) | 1 (0.9) | 3 (2.8) | 0 (0.0) | 0.133 |
| Beck score at baseline | 8 (4–15) | 9 (4–15) | 9 (5–16) | 14 (8–19) | 0.641 |
| Medical history |  |  |  |  |  |
| Ischemic heart failure | 967 (50.8) | 59 (54.6) | 62 (56.9) | 8 (80.0) | 0.365 |
| Angina | 469 (24.6) | 28 (25.9) | 28 (25.7) | 2 (20.0) | 0.931 |
| CCS angina class |  |  |  |  |  |
| CCS I | 1,589 (83.5) | 93 (86.1) | 88 (80.7) | 8 (80.0) | 0.566 |
| CCS II | 164 (8.6) | 8 (7.4) | 14 (12.8) | 1 (10.0) | 0.277 |
| CCS III | 147 (7.7) | 7 (6.5) | 7 (6.4) | 1 (10.0) | 0.798 |
| Myocardial infarction | 789 (41.5) | 52 (48.1) | 52 (47.7) | 6 (60.0) | 0.188 |
| Cardiac procedures |  |  |  |  |  |
| CABG | 475 (25.0) | 33 (30.6) | 28 (25.7) | 2 (20.0) | 0.427 |
| Valve surgery | 95 (5.0) | 5 (4.6) | 15 (13.8) | 1 (10.0) | <0.001 |
| PCI | 435 (22.9) | 25 (23.1) | 32 (29.4) | 6 (60.0) | 0.295 |
| Pacemaker | 324 (17.0) | 24 (22.2) | 32 (29.4) | 5 (50.0) | <0.01 |
| Biventricular pacemaker | 345 (18.1) | 22 (20.4) | 27 (24.8) | 2 (20.0) | 0.197 |
| AICD | 757 (39.8) | 44 (40.7) | 59 (54.1) | 5 (50.0) | <0.05 |
| Arrhythmias | 748 (39.3) | 65 (60.2) | 59 (54.1) | 8 (80.0) | <0.001 |
| Hypertension | 1,128 (59.3) | 64 (59.3) | 66 (60.6) | 2 (20.0) | 0.966 |
| Hyperlipidemia | 1,239 (65.1) | 74 (68.5) | 72 (66.1) | 8 (80.0) | 0.759 |
| Peripheral vascular disease | 130 (6.8) | 6 (5.6) | 10 (9.2) | 0 (0.0) | 0.550 |
| Stroke | 184 (9.7) | 16 (14.8) | 15 (13.8) | 3 (30.0) | 0.099 |
| Diabetes | 600 (31.5) | 34 (31.5) | 44 (40.4) | 0 (0.0) | 0.156 |
| COPD | 186 (9.8) | 29 (26.9) | 17 (15.6) | 0 (0.0) | <0.001 |
| Cancer in last 5 years | 69 (3.6) | 10 (9.3) | 4 (3.7) | 1 (10.0) | <0.05 |
| Depression | 397 (20.9) | 23 (21.3) | 29 (26.6) | 3 (30.0) | 0.361 |
| Smoking |  |  |  |  |  |
| Never | 702 (36.9) | 38 (35.2) | 40 (36.7) | 3 (30.0) | 0.938 |
| Current | 331 (17.4) | 14 (13.0) | 16 (14.7) | 1 (10.0) | 0.393 |
| Alcohol use | 806 (42.4) | 48 (44.4) | 40 (36.7) | 3 (30.0) | 0.451 |
| Average of drinks per week |  |  |  |  |  |
| 0.0—0.9 | 317 (39.3) | 19 (39.6) | 17 (42.5) | 0 (0.0) | 0.923 |
| 1.0—1.9 | 199 (24.7) | 16 (33.3) | 9 (22.5) | 1 (33.3) | 0.378 |
| 2.0— | 290 (36.0) | 13 (27.1) | 14 (35.0) | 2 (66.7) | 0.456 |
| Patient no longer drinks | 294 (36.5) | 18 (37.5) | 15 (37.5) | 0 (0.0) | 0.982 |
| Hospitalizations in last 6 months |  |  |  |  |  |
| Any hospitalizations | 754 (39.6) | 42 (38.9) | 52 (47.7) | 1 (10.0) | 0.239 |
| HF hospitalizations | 504 (26.7) | 29 (26.9) | 30 (27.5) | 1 (11.1) | 0.983 |
| Non-HF hospitalizations | 322 (16.9) | 20 (18.5) | 30 (27.5) | 0 (0.0) | <0.05 |
| Renal dysfunction | 21 (1.1) | 2 (1.9) | 4 (3.7) | 0 (0.0) | 0.058 |
| Symptomatic bradycardia | 43 (2.3) | 3 (2.8) | 3 (2.8) | 0 (0.0) | 0.896 |
| Sustained ventricular tachycardia/ventri | 274 (14.4) | 19 (17.6) | 21 (19.3) | 3 (30.0) | 0.268 |
| Claudication | 75 (3.9) | 4 (3.7) | 6 (5.5) | 0 (0.0) | 0.711 |
| Laboratory Tests |  |  |  |  |  |
| Creatinine, mg/dL | 1.2 (1.0–1.5) | 1.3 (1.1–1.7) | 1.3 (1.1–1.7) | 0.9 (0.9–1.0) | <0.01 |
| eGFR, mL/min/1.73 m^2 | 67 (52–82) | 60 (42–72) | 56 (44–76) | 68 (64–85) | <0.001 |
| Blood urea nitrogen, mg/dL | 20 (15–28) | 24 (18–34) | 23 (17–35) | 18 (17–21) | <0.05 |
| Total cholesterol, mg/dL | 162 (137–191) | 165 (138–188) | 152 (125–189) | 234 (182–250) | 0.368 |
| Low-density lipoprotein, mg/dL | 89 (70–113) | 97 (75–120) | 86 (68–116) | 117 (91–155) | 0.679 |
| High-density lipoprotein, mg/dL | 40 (33–49) | 43 (36–52) | 40 (35–46) | 52 (49–56) | 0.382 |
| Hemoglobin A1c, percent | 6.6 (5.9–7.8) | 6.4 (6.0–7.4) | 6.9 (6.4–8.2) | 5.1 (5.1–5.1) | 0.433 |
| BNP, pg/mL | 237 (104–502) | 361 (162–694) | 283 (140–949) | 136 (103–260) | 0.184 |
| Pro-BNP, pg/mL | 1183 (514–4758) | 388 (323–452) | No Data | No Data | 0.428 |
| Hemoglobin, g/dL | 14 (12–15) | 14 (12–14) | 13 (12–14) | 14 (13–14) | 0.278 |
| Medications |  |  |  |  |  |
| ACE inhibitor | 1,511 (79.4) | 80 (74.1) | 0 (0.0) | 0 (0.0) | <0.001 |
| ARB | 461 (24.2) | 32 (29.6) | 0 (0.0) | 0 (0.0) | <0.001 |
| ACE inhibitor or ARB | 1,903 (100.0) | 108 (100.0) | 0 (0.0) | 0 (0.0) | <0.001 |
| Beta blocker | 1,903 (100.0) | 0 (0.0) | 109 (100.0) | 0 (0.0) | <0.001 |
| Loop diuretic | 1,488 (78.2) | 83 (76.9) | 88 (80.7) | 5 (50.0) | 0.770 |
| Non-loop diuretic (excluding aldosterone antagonist) | 162 (8.5) | 15 (13.9) | 18 (16.5) | 0 (0.0) | <0.01 |
| Aldosterone antagonist (spiro. or epleren.) | 882 (46.3) | 44 (40.7) | 33 (30.3) | 3 (30.0) | <0.01 |
| HMG-CoA reductase inhibitor | 921 (48.4) | 39 (36.1) | 49 (45.0) | 3 (30.0) | <0.05 |
| Digoxin | 862 (45.3) | 52 (48.1) | 48 (44.0) | 2 (20.0) | 0.811 |
| Nitrates | 438 (23.0) | 34 (31.5) | 40 (36.7) | 1 (10.0) | <0.001 |
| Calcium channel blocker | 118 (6.2) | 12 (11.1) | 10 (9.2) | 1 (10.0) | 0.073 |
| Insulin | 262 (13.8) | 12 (11.1) | 24 (22.0) | 0 (0.0) | <0.05 |
| Spironolactone | 820 (43.1) | 41 (38.0) | 29 (26.6) | 1 (10.0) | <0.01 |
| Eplerenone | 63 (3.3) | 3 (2.8) | 4 (3.7) | 2 (20.0) | 0.933 |
| Beta blocker dose (carvedilol equivalent), mg | 40 (25–50) | 0 (0–0) | 25 (13–50) | 0 (0–0) | <0.01 |
| Loop diuretic dose (furosemide equivalent), mg | 40 (20–80) | 40 (12–80) | 60 (20–120) | 10 (0–35) | <0.05 |
| 6-Minute Walk Test |  |  |  |  |  |
| Patient able to walk | 1,860 (97.7) | 108 (100.0) | 107 (98.2) | 9 (90.0) | 0.278 |
| Symptomatic (e.g., angina, light-headedness, and syncope) | 212 (11.1) | 14 (13.0) | 11 (10.1) | 1 (10.0) | 0.787 |
| Total distance walked, meter | 375 (301–437) | 347 (276–439) | 329 (251–410) | 366 (322–405) | <0.01 |
| Work done during 6-minute walk, kJ | 324 (249–412) | 290 (215–374) | 279 (208–370) | 242 (183–382) | <0.001 |
| Kansas City Cardiomyopathy Questionnaire (KCCQ) |  |  |  |  |  |
| KCCQ overall summary score | 68 (52–83) | 64 (45–82) | 67 (48–81) | 65 (48–76) | 0.098 |
| KCCQ clinical summary score | 75 (59–88) | 72 (49–84) | 71 (54–83) | 74 (62–80) | <0.01 |
| KCCQ physical limitation score | 75 (54–88) | 67 (46–83) | 67 (50–83) | 68 (64–82) | <0.05 |
| KCCQ social limitation score | 67 (42–88) | 56 (38–86) | 63 (40–83) | 44 (31–75) | 0.518 |
| KCCQ total symptom score | 77 (60–91) | 73 (51–88) | 73 (53–85) | 76 (60–82) | <0.01 |
| KCCQ symptom stability score | 50 (50–50) | 50 (50–50) | 50 (50–50) | 50 (50–50) | 0.387 |
| KCCQ symptom frequency score | 77 (58–92) | 71 (54–88) | 72 (52–86) | 73 (60–81) | <0.05 |
| KCCQ symptom burden score | 83 (67–92) | 75 (56–92) | 75 (58–92) | 75 (60–90) | <0.01 |
| KCCQ self-efficacy score | 88 (75–100) | 88 (75–100) | 88 (75–100) | 82 (53–97) | 0.884 |
| KCCQ quality of life score | 58 (42–83) | 58 (42–75) | 58 (42–75) | 58 (27–83) | 0.377 |
| EuroQol Questionnaire |  |  |  |  |  |
| EuroQol thermometer response | 70 (50–80) | 68 (50–80) | 70 (52–80) | 60 (41–74) | 0.509 |
| CPX Test |  |  |  |  |  |
| Rest ECG |  |  |  |  |  |
| Normal | 807 (42.4) | 38 (35.2) | 40 (36.7) | 4 (40.0) | 0.183 |
| LBBB | 308 (16.2) | 17 (15.7) | 13 (11.9) | 0 (0.0) | 0.497 |
| RBBB | 64 (3.4) | 5 (4.6) | 6 (5.5) | 2 (20.0) | 0.410 |
| IVCD | 242 (12.7) | 14 (13.0) | 13 (11.9) | 1 (10.0) | 0.968 |
| Paced | 435 (22.9) | 31 (28.7) | 34 (31.2) | 3 (30.0) | 0.060 |
| Heart rate (CPX), bpm |  |  |  |  |  |
| At peak exercise | 120 (104–134) | 123 (107–141) | 116 (96–136) | 138 (129–147) | 0.131 |
| Resting | 70 (62–78) | 75 (67–81) | 72 (65–82) | 80 (66–86) | <0.001 |
| Reserve (peak — rest) | 48 (35–62) | 48 (29–65) | 43 (24–56) | 54 (47–73) | <0.01 |
| Borg RPE score at peak exercise | 17 (15–19) | 17 (15–18) | 17 (15–18) | 15 (15–17) | 0.166 |
| Parameters |  |  |  |  |  |
| Exercise duration, min | 9.8 (7.0–12.0) | 8.8 (6.0–11.1) | 8.5 (5.3–11.2) | 11 (10–15) | <0.01 |
| Peak VO2 (oxygen consumption), mL/kg/min | 14 (12–18) | 14 (11–18) | 13 (11–16) | 18 (17–22) | <0.05 |
| Absolute peak VO2, mL/min | 1326 (990–1701) | 1238 (982–1634) | 1193 (924–1495) | 1515 (1225–1630) | <0.01 |
| Weber class |  |  |  |  |  |
| A | 253 (13.3) | 12 (11.1) | 13 (11.9) | 4 (40.0) | 0.752 |
| B | 435 (22.9) | 28 (25.9) | 19 (17.4) | 4 (40.0) | 0.303 |
| C | 898 (47.2) | 53 (49.1) | 53 (48.6) | 2 (20.0) | 0.896 |
| D | 267 (14.0) | 13 (12.0) | 24 (22.0) | 0 (0.0) | 0.054 |
| VO2 at ventilatory threshold, mL/kg/min | 11 (9–12) | 10 (9–13) | 10 (8–12) | 12 (11–13) | 0.381 |
| VT (ventilatory threshold) percent max VO2 | 0.7 (0.6–0.8) | 0.7 (0.6–0.8) | 0.7 (0.7–0.8) | 0.7 (0.6–0.7) | <0.05 |
| Peak RER (respiratory exchange ratio) | 1.1 (1.0–1.2) | 1.1 (1.0–1.1) | 1.1 (1.0–1.2) | 1.1 (1.1–1.1) | 0.284 |
| Time to RER = 1.0, min | 7.8 (5.8–9.8) | 7.0 (4.6–9.6) | 6.7 (4.5–9.3) | 7.8 (6.4–10.3) | <0.01 |
| VeVCO2 slope | 33 (28–38) | 32 (28–38) | 33 (29–39) | 29 (28–31) | 0.226 |
| Heart rate at the end of WL 2, bpm | 91 (81–102) | 96 (88–110) | 92 (81–106) | 94 (84–104) | <0.001 |
| Peak oxygen pulse, mL/kg | 0.1 (0.1–0.1) | 0.1 (0.1–0.1) | 0.1 (0.1–0.1) | 0.1 (0.1–0.1) | 0.111 |
| Oxygen uptake efficiency slope |  |  |  |  |  |
| Entire CPX test | 1644 (1224–2081) | 1625 (1298–2022) | 1473 (1179–1913) | 1774 (1518–2223) | <0.05 |
| The first 6 minutes of CPX test | 1691 (1329–2080) | 1699 (1337–2103) | 1585 (1328–2053) | 1756 (1356–1929) | 0.736 |
| While patient's RER = 1.0 | 1688 (1278–2105) | 1720 (1312–2245) | 1529 (1366–1924) | 1818 (1624–1968) | 0.315 |
| Echocardiography |  |  |  |  |  |
| E velocity, cm/sec | 72 (56–90) | 69 (54–87) | 77 (54–100) | 69 (46–85) | 0.317 |
| A velocity, cm/sec | 65 (47–82) | 66 (45–92) | 56 (34–81) | 75 (65–81) | 0.051 |
| e´ velocity, cm/sec |  |  |  |  |  |
| Septal annulus | 5.8 (4.2–9.4) | 5.9 (4.2–9.5) | 4.9 (4.0–6.3) | 5.8 (5.0–6.9) | 0.786 |
| Lateral annulus | 8.4 (5.9–12.6) | 7.7 (5.5–11.9) | 8.0 (5.2–12.3) | 6.9 (4.4–8.9) | 0.316 |
| Average | 6.7 (5.1–9.0) | 6.6 (5.1–8.4) | 6.2 (4.6–8.2) | 6.5 (5.6–7.2) | 0.289 |
| E/A ratio | 1.1 (0.7–1.7) | 1.0 (0.7–1.7) | 1.3 (0.8–2.7) | 0.8 (0.6–1.4) | <0.05 |
| E/e´ ratio | 11 (7–15) | 10.3 (7.1–13.1) | 12 (8–19) | 10 (10–12) | 0.143 |
| Valsalva Maneuver |  |  |  |  |  |
| E velocity, cm/sec | 59 (43–80) | 58 (45–81) | 70 (56–83) | 33 (28–65) | 0.593 |
| A velocity, cm/sec | 63 (47–79) | 58 (45–74) | 61 (48–83) | 85 (78–86) | 0.952 |
| E/A ratio | 0.9 (0.7–1.4) | 1.0 (0.6–1.6) | 1.1 (0.7–1.6) | 0.4 (0.3–0.9) | 0.679 |
| E-wave Deceleration time, msec | 188 (149–233) | 180 (147–224) | 172 (136–220) | 165 (156–199) | 0.165 |
| Left atrial dimension, cm | 4.4 (3.9–4.9) | 4.5 (4.1–5.1) | 4.4 (3.9–5.1) | 4.8 (4.5–5.2) | 0.452 |
| Left ventricular diastolic dimension, cm | 6.6 (5.9–7.4) | 6.8 (6.0–7.5) | 6.5 (5.7–7.5) | 7.6 (6.6–8.4) | 0.360 |
| Left ventricular mass, g | 293 (211–388) | 307 (225–368) | 299 (226–397) | 335 (256–395) | 0.462 |
| Mitral regurgitation, percent | 17 (10–28) | 19 (11–32) | 16 (9–30) | 11 (8–17) | 0.055 |

Heterogeneity between groups were tested using ANOVA or the Kruskal-Wallis test for continuous variables and the chi-square test for categorical variables. BB Beta-blocker; RASI Angiotensin-converting enzyme inhibitor and/or angiotensin II receptor blocker.

#### Table S3. Results of Cox Regression Analysis for All-Cause Death

| **Parameter** | **Label** | **Object, n (%)** | **Control, n (%)** | **Coefficient** | **P value** | **HR (95%CI)** | **Bootstrap 95%CI** | **Rate Selected in Backward Elimination** |
| --- | --- | --- | --- | --- | --- | --- | --- | --- |
| **Main effects of exercise** |  |  |  |  |  |  |  |  |
| exercise__rasi1_beta1 | Exercise effect in patients treated with both RASI and BB at baseline | – | – | -0.15 | 0.225 | 0.86  (0.68 – 1.10) | 0.86  (0.75 – 0.97) | 100% (Fixed) |
| exercise__rasi1_beta0 | Exercise effect in patients treated with RASI but not BB at baseline | – | – | 1.52 | 0.016 | 4.56  (1.32 – 15.6) | 4.60  (3.08 – 7.08) | 100% (Fixed) |
| exercise__rasi0_beta1 | Exercise effect in patients treated with BB but not RASI at baseline | – | – | -0.45 | 0.249 | 0.64  (0.30 – 1.37) | 0.63  (0.39 – 0.97) | 100% (Fixed) |
| exercise__rasi0_beta0 | Exercise effect in patients treated with niether RASI nor BB at baseline | – | – | 0 fixed | – | – | – | – |
| **Interaction effects on exercise** |  |  |  |  |  |  |  |  |
| exercise * bmi_ge_25 | Interaction effect between exercise and bmi_ge_25 | – | – | 0.50 | 0.041 | 1.64  (1.02 – 2.64) | 1.64  (1.01 – 2.75) | 67.2% |
| exercise * ecgcond3_eq_1 | Interaction effect between exercise and ecgcond3_eq_1 | – | – | -0.47 | 0.041 | 0.62  (0.40 – 0.98) | 0.62  (0.38 – 0.96) | 63.2% |
| exercise * pulsepr_ge_30 | Interaction effect between exercise and pulsepr_ge_30 | – | – | -0.84 | 0.008 | 0.43  (0.23 – 0.81) | 0.42  (0.23 – 0.81) | 75.6% |
| exercise * hemogloc_ge_12p4 | Interaction effect between exercise and hemogloc_ge_12p4 | – | – | -0.86 | <0.001 | 0.42  (0.26 – 0.70) | 0.41  (0.24 – 0.72) | 95.3% |
| exercise * stroke_eq_1 | Interaction effect between exercise and stroke_eq_1 | – | – | -1.13 | 0.002 | 0.32  (0.16 – 0.65) | 0.31  (0.15 – 0.65) | 92.1% |
| **Other main effects** |  |  |  |  |  |  |  |  |
| **Medication at baseline** |  |  |  |  |  |  |  |  |
| acearb1_betab1 | Treated with both RASI and BB at baseline | 1,888 (89.4%) | – | reference | – | – | – | 100% (Fixed) |
| acearb1_betab0 | Treated with RASI but not BB at baseline | 107 (5.07%) | – | -1.13 | 0.054 | 0.32  (0.10 – 1.02) | 0.32  (0.25 – 0.42) | 100% (Fixed) |
| acearb0_betab1 | Treated with BB but not RASI at baseline | 107 (5.07%) | – | 0.68 | 0.004 | 1.97  (1.24 – 3.13) | 2.00  (1.54 – 2.76) | 100% (Fixed) |
| acearb0_betab0 | Treated with niether RASI nor BB at baseline | 10 (0.47%) | – | 0.66 | 0.357 | 1.94  (0.47 – 7.95) | 1.98  (1.36 – 2.98) | 100% (Fixed) |
| **Included in interactions with exercise** |  |  |  |  |  |  |  |  |
| bmi_ge_25 | Body mass index ≥25 kg/m^2 | 1,704 (80.0%) | 426 (20.0%) | -0.57 | <0.001 | 0.57  (0.41 – 0.78) | 0.57  (0.41 – 0.81) | 92.2% |
| ecgcond3_eq_1 | Normal ECG | 889 (41.7%) | 1,241 (58.3%) | 0.17 | 0.287 | 1.19  (0.87 – 1.63) | 1.18  (0.86 – 1.63) | 65.7% |
| pulsepr_ge_30 | Pulse pressure ≥30 mmHg | 1,899 (89.2%) | 231 (10.8%) | 0.26 | 0.293 | 1.29  (0.80 – 2.10) | 1.32  (0.81 – 2.24) | 77.9% |
| hemogloc_ge_12p4 | Hemoglobin ≥12.4 g/dL | 1,720 (80.8%) | 410 (19.2%) | 0.24 | 0.211 | 1.27  (0.87 – 1.86) | 1.28  (0.88 – 1.97) | 95.5% |
| stroke_eq_1 | History of stroke | 218 (10.2%) | 1,912 (89.8%) | 0.45 | 0.027 | 1.57  (1.05 – 2.35) | 1.59  (1.01 – 2.44) | 92.7% |
| **Others** |  |  |  |  |  |  |  |  |
| vevco2_ge_34 | Ve/VCO2 slope (CPX test) ≥34 | 926 (43.5%) | 1,204 (56.5%) | 0.53 | <0.001 | 1.70  (1.35 – 2.15) | 1.72  (1.32 – 2.17) | 88.7% |
| valvsurg_eq_1 | History of prior valve surgery | 116 (5.45%) | 2,014 (94.6%) | 0.50 | 0.004 | 1.65  (1.18 – 2.31) | 1.67  (1.16 – 2.30) | 85.6% |
| male_eq_1 | Mele sex | 1,531 (71.9%) | 599 (28.1%) | 0.42 | 0.002 | 1.52  (1.16 – 1.99) | 1.52  (1.17 – 2.02) | 97.2% |
| bestlvef_ge_20 | Best available baseline LVEF ≥20 % | 1,616 (75.9%) | 514 (24.1%) | -0.43 | <0.001 | 0.65  (0.52 – 0.82) | 0.65  (0.52 – 0.83) | 81.4% |
| egfr_ge_65 | eGFR ≥65 mL/min/1.73 m^2 | 1,162 (54.6%) | 968 (45.4%) | -0.56 | <0.001 | 0.57  (0.45 – 0.72) | 0.57  (0.43 – 0.71) | 99.1% |
| hrpeakex_ge_100 | HR at peak exercise (CPX test) ≥100 bpm | 1,725 (81.0%) | 405 (19.0%) | -0.68 | <0.001 | 0.51  (0.40 – 0.64) | 0.50  (0.39 – 0.64) | 87.7% |

BB Beta-blocker; CI Confidence interval; CPX Cardiopulmonary exercise testing; ECG Electrocardiogram; eGFR estimated glomerular filtration rate; HR Hazard ratio; HR Heart rate; LVEF Left ventricle ejection fraction; RASI Angiotensin-converting enzyme inhibitor and/or angiotensin II receptor blocker; VCO2 Carbon dioxide output; Ve Ventilatory equivalent.

Linear predictors (e.q., ln(HR)) are calculated according to the following equation:

Linear predictors (e.q., ln (HR)) are calculated according to the following equation:

ln(HR) = –0.15 * exercise__rasi1_beta1[=1.00 if yes else 0.00] + 1.52 * exercise__rasi1_beta0[=1.00 if yes else 0.00] – 0.45 * exercise__rasi0_beta1[=1.00 if yes else 0.00] – 1.13 * rasi1_beta0[=0.95 if yes else –0.05] + 0.68 * rasi0_beta1[=0.95 if yes else –0.05] + 0.66 * rasi0_beta0[=1.00 if yes else –0.00] – 0.42 * male_eq_0[=0.72 if yes else –0.28] + 0.57 * bmi_lt_25[=0.80 if yes else –0.20] + 0.56 * egfr_lt_65[=0.55 if yes else –0.45] + 0.43 * bestlvef_lt_20[=0.76 if yes else –0.24] + 0.53 * vevco2_ge_34[=0.57 if yes else –0.43] + 0.68 * hrpeakex_lt_100[=0.81 if yes else –0.19] – 0.24 * hemogloc_lt_12p4[=0.81 if yes else –0.19] + 0.50 * valvsurg_eq_1[=0.95 if yes else –0.05] + 0.45 * stroke_eq_1[=0.90 if yes else –0.10] – 0.26 * pulsepr_lt_30[=0.89 if yes else –0.11] + 0.17 * ecgcond3_eq_1[=0.58 if yes else –0.42] – 0.50 * exercise[=1.00 if yes else 0.00] * bmi_lt_25[=0.80 if yes else –0.20] + 0.84 * exercise[=1.00 if yes else 0.00] * pulsepr_lt_30[=0.89 if yes else –0.11] – 0.47 * exercise[=1.00 if yes else 0.00] * ecgcond3_eq_1[=0.58 if yes else –0.42] + 0.86 * exercise[=1.00 if yes else 0.00] * hemogloc_lt_12p4[=0.81 if yes else –0.19] – 1.13 * exercise[=1.00 if yes else 0.00] * stroke_eq_1[=0.90 if yes else –0.10]

#### Table S4. Results of Cox Regression Analysis for All-Cause Death or All-Cause Hospitalization

| **Parameter** | **Label** | **Object, n (%)** | **Control, n (%)** | **Coefficient** | **P value** | **HR (95%CI)** | **Bootstrap 95%CI** | **Rate Selected in Backward Elimination** |
| --- | --- | --- | --- | --- | --- | --- | --- | --- |
| **Main effects of exercise** |  |  |  |  |  |  |  |  |
| exercise__rasi1_beta1 | Exercise effect in patients treated with both RASI and BB at baseline | – | – | -0.11 | 0.056 | 0.90  (0.80 – 1.00) | 0.90  (0.83 – 0.97) | 100% (Fixed) |
| exercise__rasi1_beta0 | Exercise effect in patients treated with RASI but not BB at baseline | – | – | -0.02 | 0.923 | 0.98  (0.62 – 1.53) | 0.98  (0.69 – 1.37) | 100% (Fixed) |
| exercise__rasi0_beta1 | Exercise effect in patients treated with BB but not RASI at baseline | – | – | -0.58 | 0.015 | 0.56  (0.35 – 0.89) | 0.55  (0.40 – 0.79) | 100% (Fixed) |
| exercise__rasi0_beta0 | Exercise effect in patients treated with niether RASI nor BB at baseline | – | – | 0 fixed | – | – | – | – |
| **Interaction effects on exercise** |  |  |  |  |  |  |  |  |
| exercise * arr_eq_1 | Interaction between exercise and arr_eq_1 | 449 (51.0%) | 431 (49.0%) | 0.44 | <0.001 | 1.55  (1.26 – 1.92) | 1.57  (1.25 – 1.93) | 95.5% |
| exercise * dbp_ge_60 | Interaction between exercise and dbp_ge_60 | 922 (49.6%) | 935 (50.4%) | 0.43 | 0.006 | 1.53  (1.13 – 2.08) | 1.54  (1.15 – 2.10) | 84.7% |
| exercise * vtpctvo2_ge_0p7 | Interaction between exercise and vtpctvo2_ge_0p7 | 628 (49.4%) | 642 (50.6%) | -0.31 | 0.007 | 0.74  (0.59 – 0.92) | 0.73  (0.59 – 0.91) | 80.5% |
| exercise * stroke_eq_1 | Interaction between exercise and stroke_eq_1 | 105 (48.2%) | 113 (51.8%) | -0.44 | 0.009 | 0.64  (0.46 – 0.90) | 0.64  (0.46 – 0.90) | 65.0% |
| exercise * pulsepr_ge_30 | Interaction between exercise and pulsepr_ge_30 | 943 (49.7%) | 956 (50.3%) | -0.67 | <0.001 | 0.51  (0.37 – 0.71) | 0.51  (0.37 – 0.70) | 97.4% |
| **Other main effects** |  |  |  |  |  |  |  |  |
| **Medication at baseline** |  |  |  |  |  |  |  |  |
| acearb1_betab1 | Treated with both RASI and BB at baseline | 1,888 (89.4%) | – | reference | . | . | . | 100% (Fixed) |
| acearb1_betab0 | Treated with RASI but not BB at baseline | 107 (5.07%) | – | -0.02 | 0.916 | 0.98  (0.70 – 1.37) | 0.99  (0.79 – 1.27) | 100% (Fixed) |
| acearb0_betab1 | Treated with BB but not RASI at baseline | 107 (5.07%) | – | 0.38 | 0.008 | 1.47  (1.11 – 1.95) | 1.49  (1.16 – 1.87) | 100% (Fixed) |
| acearb0_betab0 | Treated with niether RASI nor BB at baseline | 10 (0.47%) | – | 0.64 | 0.076 | 1.90  (0.94 – 3.88) | 1.88  (1.08 – 4.52) | 100% (Fixed) |
| **Included in interactions with exercise** |  |  |  |  |  |  |  |  |
| arr_eq_1 | History of arrhythmias | 880 (41.3%) | 1,250 (58.7%) | 0.05 | 0.489 | 1.05  (0.91 – 1.23) | 1.05  (0.91 – 1.22) | 99.9% |
| dbp_ge_60 | Diastolic blood pressure ≥60 mmHg | 1,857 (87.2%) | 273 (12.8%) | -0.19 | 0.082 | 0.82  (0.66 – 1.03) | 0.82  (0.67 – 1.01) | 86.0% |
| vtpctvo2_ge_0p7 | VO2 at VT / peak VO2 (CPX test) ≥0.7 | 1,270 (59.6%) | 860 (40.4%) | 0.37 | <0.001 | 1.45  (1.23 – 1.71) | 1.45  (1.23 – 1.71) | 98.7% |
| stroke_eq_1 | History of stroke | 218 (10.2%) | 1,912 (89.8%) | 0.31 | 0.006 | 1.37  (1.10 – 1.71) | 1.37  (1.10 – 1.70) | 69.2% |
| pulsepr_ge_30 | Pulse pressure ≥30 mmHg | 1,899 (89.2%) | 231 (10.8%) | 0.16 | 0.201 | 1.17  (0.92 – 1.49) | 1.18  (0.91 – 1.49) | 98.2% |
| **Others** |  |  |  |  |  |  |  |  |
| sixmwlks_eq_1 | Six minute walk: symptomatic?, Yes | 238 (11.2%) | 1,892 (88.8%) | 0.28 | <0.001 | 1.32  (1.13 – 1.55) | 1.33  (1.13 – 1.57) | 90.5% |
| nitrate_eq_1 | Use of nitrates at baseline | 513 (24.1%) | 1,617 (75.9%) | 0.19 | 0.002 | 1.21  (1.07 – 1.36) | 1.21  (1.07 – 1.37) | 82.2% |
| dbp_ge_70 | Diastolic blood pressure ≥70 mmHg | 1,162 (54.6%) | 968 (45.4%) | -0.14 | 0.018 | 0.87  (0.77 – 0.98) | 0.86  (0.77 – 0.97) | 89.4% |
| sixmwlkd_ge_370 | Six-minute walk distance, subjects able to walk ≥370 meters | 1,079 (50.7%) | 1,051 (49.3%) | -0.18 | 0.003 | 0.83  (0.74 – 0.94) | 0.83  (0.74 – 0.93) | 94.8% |
| white_eq_1 | Race, White | 1,354 (63.6%) | 776 (36.4%) | -0.18 | 0.004 | 0.84  (0.75 – 0.95) | 0.84  (0.74 – 0.94) | 88.3% |
| ecgcond3_eq_1 | Normal ECG | 889 (41.7%) | 1,241 (58.3%) | -0.21 | <0.001 | 0.81  (0.72 – 0.91) | 0.81  (0.72 – 0.91) | 87.0% |
| bestlvef_ge_20 | Best available baseline LVEF ≥20 % | 1,616 (75.9%) | 514 (24.1%) | -0.22 | <0.001 | 0.80  (0.71 – 0.90) | 0.80  (0.71 – 0.90) | 83.9% |
| pko2puls_ge_0p10 | Peak oxygen pulse (CPX test) ≥0.10 mL/kg | 1,616 (75.9%) | 514 (24.1%) | -0.25 | <0.001 | 0.78  (0.69 – 0.89) | 0.78  (0.68 – 0.89) | 95.8% |
| kccqosb_ge_50 | KCCQ overall summary score at baseline ≥50 | 1,639 (76.9%) | 491 (23.1%) | -0.28 | <0.001 | 0.76  (0.66 – 0.86) | 0.75  (0.66 – 0.86) | 91.8% |
| cpxdur_ge_12 | Exercise duration (CPX test) ≥12 minutes | 566 (26.6%) | 1,564 (73.4%) | -0.28 | <0.001 | 0.76  (0.65 – 0.88) | 0.76  (0.65 – 0.87) | 82.3% |
| egfr_ge_50 | eGFR ≥50 mL/min/1.73 m^2 | 1,660 (77.9%) | 470 (22.1%) | -0.32 | <0.001 | 0.73  (0.64 – 0.82) | 0.72  (0.64 – 0.82) | 82.1% |
| kccqssb_ge_50 | KCCQ symptom stability score at baseline ≥50 | 1,965 (92.3%) | 165 (7.75%) | -0.48 | <0.001 | 0.62  (0.52 – 0.75) | 0.61  (0.50 – 0.75) | 99.4% |

BB Beta-blocker; CI Confidence interval; CPX Cardiopulmonary exercise testing; ECG Electrocardiogram; eGFR estimated glomerular filtration rate; HR Hazard ratio; KCCQ Kansas City Cardiomyopathy Questionnaire; LVEF Left ventricle ejection fraction; RASI Angiotensin-converting enzyme inhibitor and/or angiotensin II receptor blocker; VT Ventilation Threshold; VO2 Oxygen uptake.

Linear predictors (e.q., ln (HR)) are calculated according to the following equation:

ln(HR) = –0.11 * exercise__rasi1_beta1[=1.00 if yes else 0.00] – 0.02 * exercise__rasi1_beta0[=1.00 if yes else 0.00] – 0.58 * exercise__rasi0_beta1[=1.00 if yes else 0.00] – 0.02 * rasi1_beta0[=0.95 if yes else –0.05] + 0.38 * rasi0_beta1[=0.95 if yes else –0.05] + 0.64 * rasi0_beta0[=1.00 if yes else –0.00] + 0.18 * white_eq_0[=0.64 if yes else –0.36] – 0.16 * pulsepr_lt_30[=0.89 if yes else –0.11] + 0.19 * dbp_lt_60[=0.87 if yes else –0.13] + 0.14 * dbp_lt_70[=0.55 if yes else –0.45] + 0.32 * egfr_lt_50[=0.78 if yes else –0.22] + 0.22 * bestlvef_lt_20[=0.76 if yes else –0.24] – 0.21 * ecgcond3_eq_1[=0.58 if yes else –0.42] – 0.28 * cpxdur_ge_12[=0.73 if yes else –0.27] – 0.37 * vtpctvo2_lt_0p7[=0.60 if yes else –0.40] + 0.25 * pko2puls_lt_0p10[=0.76 if yes else –0.24] + 0.28 * sixmwlks_eq_1[=0.89 if yes else –0.11] + 0.18 * sixmwlkd_lt_370[=0.51 if yes else –0.49] + 0.05 * arr_eq_1[=0.59 if yes else –0.41] + 0.19 * nitrate_eq_1[=0.76 if yes else –0.24] + 0.28 * kccqosb_lt_50[=0.77 if yes else –0.23] + 0.48 * kccqssb_lt_50[=0.92 if yes else –0.08] + 0.31 * stroke_eq_1[=0.90 if yes else –0.10] + 0.67 * exercise[=1.00 if yes else 0.00] * pulsepr_lt_30[=0.89 if yes else –0.11] – 0.43 * exercise[=1.00 if yes else 0.00] * dbp_lt_60[=0.87 if yes else –0.13] + 0.31 * exercise[=1.00 if yes else 0.00] * vtpctvo2_lt_0p7[=0.60 if yes else –0.40] – 0.44 * exercise[=1.00 if yes else 0.00] * stroke_eq_1[=0.90 if yes else –0.10] + 0.44 * exercise[=1.00 if yes else 0.00] * arr_eq_1[=0.59 if yes else –0.41]

#### Table S5. Sensitivity Analysis for All-Cause Mortality

|  | **final model** | **experiment 1** | **experiment 2** |
| --- | --- | --- | --- |
| **exercise** | – | 0.89 (0.70—1.12) | 0.96 (0.78—1.18) |
| **exercise__rasi1_beta1** | 0.86 (0.75—0.97) | – | – |
| **exercise__rasi1_beta0** | 4.60 (3.08—7.08) | – | – |
| **exercise__rasi0_beta1** | 0.63 (0.39—0.97) | – | – |
| **exercise__rasi0_beta0** | 0 fixed | – | – |
| **rasi1_beta1** | 0 fixed | 0 fixed | 0 fixed |
| **rasi1_beta0** | 0.32 (0.25—0.42) | 1.03 (0.66—1.52) | 1.06 (0.67—1.55) |
| **rasi0_beta1** | 2.00 (1.54—2.76) | 1.79 (1.19—2.68) | 1.74 (1.18—2.58) |
| **rasi0_beta0** | 1.98 (1.36—2.98) | 2.24 (0.00—4.20) | 1.64 (0.00—3.31) |
| **male_eq_1** | 1.52 (1.17—2.02) | 1.53 (1.17—2.08) | 1.54 (1.19—2.08) |
| **bmi_ge_25** | 0.57 (0.41—0.81) | 0.57 (0.41—0.80) | 0.69 (0.55—0.90) |
| **egfr_ge_65** | 0.57 (0.43—0.71) | 0.56 (0.44—0.69) | 0.56 (0.45—0.69) |
| **bestlvef_ge_20** | 0.65 (0.52—0.83) | 0.64 (0.50—0.80) | 0.63 (0.50—0.79) |
| **vevco2_ge_34** | 1.72 (1.32—2.17) | 1.70 (1.37—2.13) | 1.68 (1.36—2.10) |
| **hrpeakex_ge_100** | 0.50 (0.39—0.64) | 0.50 (0.39—0.65) | 0.53 (0.42—0.68) |
| **hemogloc_ge_12p4** | 1.28 (0.88—1.97) | 1.24 (0.88—1.88) | 0.84 (0.64—1.09) |
| **valvsurg_eq_1** | 1.67 (1.16—2.30) | 1.62 (1.12—2.28) | 1.67 (1.20—2.35) |
| **stroke_eq_1** | 1.59 (1.01—2.44) | 1.60 (0.99—2.39) | 1.04 (0.70—1.46) |
| **pulsepr_ge_30** | 1.32 (0.81—2.24) | 1.26 (0.80—2.11) | 0.87 (0.62—1.21) |
| **ecgcond3_eq_1** | 1.18 (0.86—1.63) | 1.18 (0.87—1.70) | 0.96 (0.76—1.22) |
| **exercise * bmi_ge_25** | 1.64 (1.01—2.75) | 1.63 (0.96—2.81) | – |
| **exercise * pulsepr_ge_30** | 0.42 (0.23—0.81) | 0.43 (0.20—0.81) | – |
| **exercise * ecgcond3_eq_1** | 0.62 (0.38—0.96) | 0.61 (0.37—0.96) | – |
| **exercise * hemogloc_ge_12p4** | 0.41 (0.24—0.72) | 0.43 (0.24—0.69) | – |
| **exercise * stroke_eq_1** | 0.31 (0.15—0.65) | 0.32 (0.13—0.68) | – |

#### Table S6. Sensitivity analysis for All-Cause Mortality or All-Cause Hospitalization

|  | **final model** | **experiment 1** | **experiment 2** |
| --- | --- | --- | --- |
| **exercise** | – | 0.88 (0.82—0.94) | 0.88 (0.82—0.94) |
| **exercise__rasi1_beta1** | 0.90 (0.83—0.97) | – | – |
| **exercise__rasi1_beta0** | 0.98 (0.69—1.37) | – | – |
| **exercise__rasi0_beta1** | 0.55 (0.40—0.79) | – | – |
| **exercise__rasi0_beta0** | 0 fixed | – | – |
| **rasi1_beta1** | 0 fixed | 0 fixed | 0 fixed |
| **rasi1_beta0** | 0.99 (0.79—1.27) | 1.03 (0.87—1.24) | 1.04 (0.90—1.23) |
| **rasi0_beta1** | 1.49 (1.16—1.87) | 1.22 (1.03—1.47) | 1.23 (1.06—1.47) |
| **rasi0_beta0** | 1.88 (1.08—4.52) | 2.00 (1.16—4.76) | 1.96 (1.10—5.05) |
| **white_eq_1** | 0.84 (0.74—0.94) | 0.83 (0.74—0.94) | 0.85 (0.76—0.96) |
| **pulsepr_ge_30** | 1.18 (0.91—1.49) | 1.16 (0.89—1.46) | 0.85 (0.72—1.00) |
| **dbp_ge_60** | 0.82 (0.67—1.01) | 0.83 (0.67—1.02) | 1.02 (0.86—1.20) |
| **dbp_ge_70** | 0.86 (0.77—0.97) | 0.86 (0.77—0.97) | 0.86 (0.77—0.97) |
| **egfr_ge_50** | 0.72 (0.64—0.82) | 0.73 (0.64—0.82) | 0.74 (0.65—0.84) |
| **bestlvef_ge_20** | 0.80 (0.71—0.90) | 0.79 (0.71—0.89) | 0.78 (0.70—0.88) |
| **ecgcond3_eq_1** | 0.81 (0.72—0.91) | 0.81 (0.72—0.90) | 0.82 (0.73—0.92) |
| **cpxdur_ge_12** | 0.76 (0.65—0.87) | 0.75 (0.66—0.87) | 0.77 (0.67—0.89) |
| **vtpctvo2_ge_0p7** | 1.45 (1.23—1.71) | 1.46 (1.24—1.72) | 1.25 (1.10—1.42) |
| **pko2puls_ge_0p10** | 0.78 (0.68—0.89) | 0.78 (0.69—0.89) | 0.77 (0.68—0.89) |
| **sixmwlks_eq_1** | 1.33 (1.13—1.57) | 1.31 (1.11—1.55) | 1.31 (1.10—1.56) |
| **sixmwlkd_ge_370** | 0.83 (0.74—0.93) | 0.83 (0.75—0.93) | 0.83 (0.75—0.93) |
| **arr_eq_1** | 1.05 (0.91—1.22) | 1.05 (0.91—1.23) | 1.30 (1.17—1.44) |
| **nitrate_eq_1** | 1.21 (1.07—1.37) | 1.21 (1.07—1.38) | 1.20 (1.06—1.35) |
| **kccqosb_ge_50** | 0.75 (0.66—0.86) | 0.75 (0.66—0.85) | 0.77 (0.67—0.87) |
| **kccqssb_ge_50** | 0.61 (0.50—0.75) | 0.62 (0.50—0.75) | 0.62 (0.50—0.76) |
| **stroke_eq_1** | 1.37 (1.10—1.70) | 1.37 (1.10—1.70) | 1.09 (0.93—1.28) |
| **exercise * pulsepr_ge_30** | 0.51 (0.37—0.70) | 0.52 (0.37—0.72) | – |
| **exercise * dbp_ge_60** | 1.54 (1.15—2.10) | 1.52 (1.13—2.07) | – |
| **exercise * vtpctvo2_ge_0p7** | 0.73 (0.59—0.91) | 0.73 (0.58—0.90) | – |
| **exercise * stroke_eq_1** | 0.64 (0.46—0.90) | 0.65 (0.46—0.92) | – |
| **exercise * arr_eq_1** | 1.57 (1.25—1.93) | 1.55 (1.24—1.90) | – |
